# Predicting emergent Dolutegravir resistance in South Africa: A modelling study

**DOI:** 10.1101/2024.07.20.24310740

**Authors:** Tom Loosli, Anthony Hauser, Johannes Josi, Nuri Han, Suzanne M Ingle, Ard van Sighem, Linda Wittkop, Janne Vehreschild, Francesca Ceccherini-Silberstein, Gary Maartens, M John Gill, Caroline A Sabin, Leigh F Johnson, Richard Lessells, Huldrych F Günthard, Matthias Egger, Roger D Kouyos

## Abstract

**Background:** In response to the rising prevalence of non-nucleoside reverse transcriptase inhibitors (NNRTIs) resistance, millions of people living with HIV (PWH) have switched to dolutegravir-based antiretroviral therapy (ART). Understanding the possible emergence of dolutegravir resistance is essential for health policy and planning. We developed a mathematical model to predict the trends of dolutegravir resistance in PWH in South Africa.

**Methods:** MARISA (Modelling Antiretroviral drug Resistance In South Africa) is a deterministic compartmental model consisting of four layers: (i) the cascade of care, (ii) disease progression, (iii) gender, and (iv) drug resistance. MARISA was calibrated to reproduce the HIV epidemic in South Africa. We assumed dolutegravir was introduced in 2020. We extended the model by including key resistance mutations observed in PWH experiencing virologic failure on dolutegravir-based ART (G118K, E138AKT, G140ACS, Q148HKNR, N155H, and R263K). Model outcomes were acquired (ADR) and transmitted drug resistance (TDR) to dolutegravir and NNRTIs stratified by duration on failing dolutegravir-based ART and under different counterfactual scenarios of switching to protease-inhibitor (PI)-based ART.

**Finding:** The model predicts that ADR will increase rapidly, from 18.5% (uncertainty range 12.5% to 25.4%) in 2023 to 46.2% (32.9% to 58.9%) in 2040. The prevalence of ADR in 2040 increased with the duration of virologic failure on dolutegravir-based ART: 18.0% (12.2% to 23.7%) for 6 months of failing ART compared to 54.8% (41.1% to 63.9%) for over 1.5 years. For TDR, the model predicts a slow but steady increase from 0.1% (0.1% to 0.2%) in 2023 to 8.8% (4.4% to 17.3%) in 2040. Transmitted NNRTI resistance will cease to increase but remain prevalent at 7.7% in 2040. Rapid resistance testing-informed switching to PI-based ART would substantially reduce both ADR and TDR.

**Interpretation:** The prevalence of dolutegravir ADR and TDR will likely increase, with the 10% threshold of TDR possibly reached by 2035, depending on monitoring and switching strategies. The increase will likely be greater in settings where resources for HIV-1 RNA monitoring and resistance testing or options for switching to alternative ART regimens are limited.

**Funding:** Swiss National Science Foundation, National Institutes of Health, UZH URPP Evolution in Action

**Research in context:** *Evidence before this study:* Dolutegravir has demonstrated high efficacy, even in individuals with compromised backbone drugs. We searched Scopus on April 15 2024, using free text words dolutegravir and resistance. We did not identify any modelling studies attempting to predict dolutegravir resistance trends in the coming years. A recent collaborative analysis of predominantly European cohort studies involving 599 people living with HIV (PWH) who underwent genotypic resistance testing at the point of dolutegravir-based treatment failure showed that the risk of dolutegravir resistance increases significantly in the presence of Nucleoside Reverse Transcriptase Inhibitor (NRTI) resistance. This is particularly concerning in settings such as South Africa, where a high proportion of individuals already exhibit NRTI resistance. Indeed, recent surveys in South Africa already hint at rapidly increasing levels of acquired dolutegravir resistance.

*Added value of this study:* This study is the first to model the likely dynamics of dolutegravir resistance in South Africa. Covering the period 2020 to 2040, it extends a previous model of antiretroviral drug resistance evolution in South Africa to dolutegravir-based ART. The results indicate that while dolutegravir resistance is currently low, it will increase at the population level, and transmitted dolutegravir resistance may exceed 10% by around 2035, depending on the duration PWH spend on failing dolutegravir-based ART. Interventions such as switching to protease-inhibitor (PI)-based ART based on genotypic resistance tests could reduce or even curb the rise of dolutegravir resistance.

*Implications of all the available evidence:* Dolutegravir resistance may undermine the success of integrase strand transfer inhibitor (INSTI)-based ART in South Africa, where the guidelines limit drug resistance testing to PWH with repeated viral load measurements above 1,000 copies/mL and evidence of good adherence. Monitoring the evolution of dolutegravir resistance at the population level is crucial to inform future changes in guidelines on drug resistance testing and switching to PI-based ART.

## Introduction

In response to the rising prevalence of drug resistance to non-nucleoside reverse transcriptase inhibitors (NNRTIs), the World Health Organization (WHO) has recommended the integrase strand transfer inhibitor (INSTI) dolutegravir as first- and second-line antiretroviral therapy (ART) for all people living with HIV (PWH) since 2018.^1^ By July 2022, dolutegravir had been adopted as the preferred first-line ART in 108 countries,^2^ including South Africa, where over 7.5 million people live with HIV.^3^ Dolutegravir-based ART has a higher genetic barrier to resistance than the previously recommended non-nucleoside reverse transcriptase inhibitor (NNRTI)-based first-line regimens.^4^ Dolutegravir-resistant HIV is currently rare in PWH on first-line ART but is observed more frequently in treatment-experienced people in trials,^5^ observational cohorts,^6,7^ and national surveys.^8–10^ A collaborative analysis of eight large cohort studies combined data from 599 individuals, primarily from Europe, who experienced viremia on dolutegravir-based ART and underwent genotypic resistance testing (GRT). At least one major or accessory INSTI drug resistance mutation (DRM) was found in 86 (14%) study participants, and 23 (3.8%) had high or intermediate levels of predicted resistance to dolutegravir.^6^

In South Africa, the current treatment guidelines recommend GRT only for people who have two or more viral load measurements exceeding 1,000 copies/mL and have been on dolutegravir-based or protease inhibitor (PI)-ART for over two years with good adherence.^11^ Consequently, individuals often remain viremic for prolonged periods.^12–14^ Furthermore, switching to alternative regimens, such as protease inhibitor (PI)-based regimens, is relatively uncommon among those on failing dolutegravir-based ART.^15^ Together, these factors may facilitate the emergence and spread of HIV drug resistance.

MARISA (Modelling Antiretroviral drug Resistance In South Africa) is a mathematical model that simulates the HIV epidemic and NNRTI resistance dynamics in South Africa.^16,17^ We updated the model to include a dimension for dolutegravir resistance, allowing us to study the emergence of acquired and transmitted resistance to dolutegravir. We aimed to predict how dolutegravir resistance levels will change over time to assess if the drug will stay effective in the long run or if resistance will rise rapidly, similar to what happened with the NNRTIs.

## Methods

### The MARISA model

Described in detail elsewhere,^16^ MARISA is a deterministic compartmental model of the HIV-1 epidemic in South Africa from 2005 to 2040 using monthly time steps. Briefly, it consists of four dimensions: i) the continuum of care from HIV infection through to the diagnosis and initiation of ART, suppressive ART (viral load < 1’000 copies per ml) and failing ART (viral load ≥ 1’000 copies per ml), and possible transfer to PI-based ART; ii) disease progression, with four CD4 count groups (> 500, 350 – 500, 200 – 349, and < 200 cells/µL); iii) gender; iv) drug resistance (appendix p3). MARISA was calibrated to reproduce the HIV epidemic in South Africa using data from the IeDEA collaboration^18^ and the literature and fitted to estimates of the demographic and epidemiological Thembisa model.^19^ We assumed the introduction of dolutegravir started in 2020. Rates of initiation and switching to dolutegravir-based ART are described elsewhere^17^ and in the appendix (p4-5).

The original implementation of MARISA treated resistance as two states (susceptible or resistant), which is appropriate for antiretroviral drugs with a low genetic barrier, such as NRTIs and NNRTIs. However, this approach is unsuitable for dolutegravir, which has a higher genetic barrier.^4^ We included the drug resistance mutations observed in the DTG RESIST study, i.e., G118K, E138AKT, G140ACS, Q148HKNR, N155H, and R263K.^6^ These six DRMs cover all mutations identified as signature DRMs for dolutegravir resistance.^20^ R263K was assumed to occur in isolation, whereas G140ACS and Q148HKNR, and G118R and E138K occurred in combination, in line with a recent review of acquired dolutegravir drug resistance mutations.^20^ The resulting genotypes (see Figure 1) were classified as susceptible, potential-low, low, intermediate, and high-level dolutegravir resistance according to the Stanford resistance algorithm.^21^ As in Hauser et al.,^16,17^ we included resistance for NRTI and NNRTI (susceptible or resistant), resulting in total in 48 drug resistance compartments. We expanded the treatment cascade, stratifying the treatment failure compartments according to the average duration on failing ART (< 6 months, 6 months to 1.5 years, > 1.5 years). Finally, we modified the model to include out-of-care dynamics, whereby individuals on failing dolutegravir-based ART may leave and re-enter care (Figure 1). See appendix p6-7 for further details.

**Figure 1:**
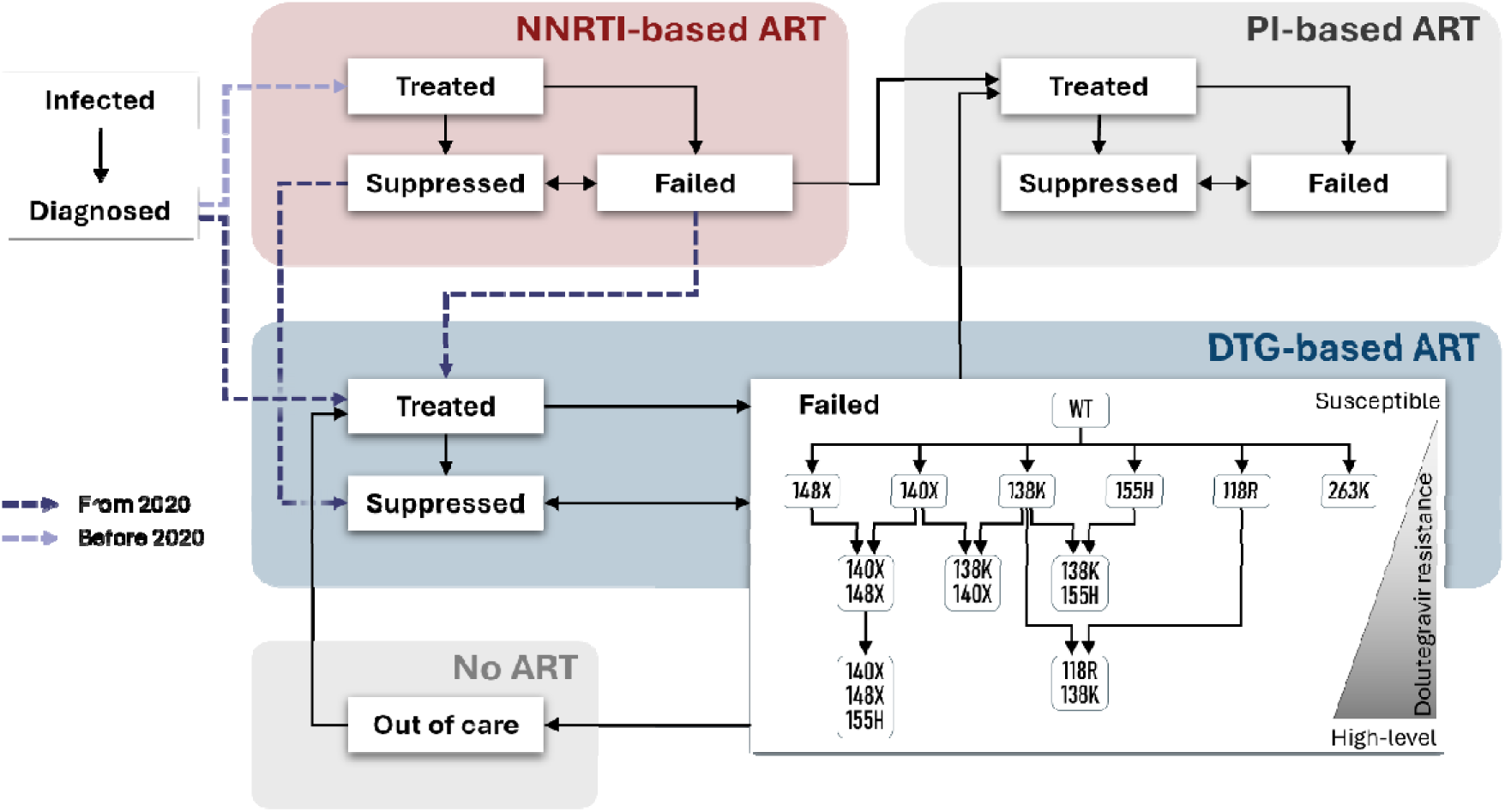
Simplified schematic overview of the adapted MARISA model. The treatment dimension, including dolutegravir resistance acquisition on failing dolutegravir-based ART, is shown. Not displayed here are the dimensions for gender and CD4 levels, NNRTI- and NRTI resistance acquisition, and mortality. Details on the model structure can be found in the appendix p3-11.

### Definitions, parameters and calibration

We defined transmitted dolutegravir resistance as the proportion of people with intermediate or high levels of dolutegravir resistance at diagnosis. Acquired drug resistance was defined as the proportion of people on a failing dolutegravir-based regimen with intermediate or high-level dolutegravir resistance.

Key model parameters included (i) acquisition and (ii) reversion rates of dolutegravir resistance mutations, (iii) the impact of NRTI resistance on dolutegravir resistance acquisition, (iv) the impact of dolutegravir resistance on dolutegravir treatment efficacy, (v) the probability of transmission dolutegravir DRMs compared to NNRTI resistance transmission, and (vi) the proportion on failing dolutegravir-based ART with detectable drug levels. Based on the DTG RESIST study,^6^ we assume that NRTI resistance increases the risk of acquiring dolutegravir DRMs upon treatment failure (appendix page 10). We estimated mutation-specific acquisition rates based on an estimated duration of 3 months of failing dolutegravir-based ART in the DTG RESIST study. The reversal of dolutegravir resistance mutations is not well documented but may occur rapidly.^22^ We assumed an average duration to reversion of two years for each mutation with HIV replicating in the absence of dolutegravir. We assumed that high- and intermediate-level dolutegravir resistance increases the treatment failure rate on dolutegravir-based ART: for high-level resistance, the impact is equal to that of high-level NNRTI resistance on NNRTI-based ART; for intermediate resistance, the effect is halved. The probability of transmitting dolutegravir resistance mutations in an HIV transmission event is unknown, but cases have been documented.^23^ We assumed an equal transmission rate for dolutegravir resistant strains as for susceptible strains, and explored decreased rates in sensitivity analyses (allowing for up to 20 times decreased transmission rates in resistant strains). The parameters included in MARISA and its calibration are described in detail in the appendix (p4-11).

To construct plausible ranges of model outcomes (predicted dolutegravir ADR and TDR) that reflect the uncertainty in the choice of parameter values, we defined an uncertainty range using pessimistic (favouring the emergence of resistance) or optimistic (impeding the emergence of resistance) parameter values in addition to our baseline parameterisation (Table 1 and appendix p12).

**Table 1:** Key dolutegravir resistance parameters. Modelled prospective scenarios include baseline parameter values, and ranges using pre-defined in- and decreased resistance parameters to derive an uncertainty interval. Other parameters used in the model can be found in the appendix p4-11. Abbreviations: DRM: drug resistance mutation.

| Parameter | Description | Baseline | Uncertainty range |  |
| --- | --- | --- | --- | --- |
|  |  |  | Pessimistic choice in parameter values | Optimistic choice in parameter values |
| $1 / r_{\text{DRM}}$ | Mean time for dolutegravir DRMs to revert (years) | 2 | 3 | 2 |
| $\alpha_{\text{NRTI} \rightarrow \text{DTG}}$ | Impact of NRTI resistance on dolutegravir DRM acquisition rates (hazard ratio vs susceptible) | 4 | 5 | 3 |
| $\alpha_{\text{impact DTG}}$ | Impact of high-level dolutegravir resistance on dolutegravir efficacy (hazard ratio vs susceptible) | 3.24 | 2 | 4 |
| $\Phi_{\text{transmission DTG}}$ | Probability of dolutegravir resistance transmission compared to NNRTI resistance transmission | Same | Same | -20% |
| $\rho_{\text{drug detect}}$ | Proportion with detectable drugs on failing dolutegravir-based ART | 0.626 | 0.814 | 0.482 |

Note: For all scenarios, mutation rates were calculated based on assumed time of 3 months on failing dolutegravir-based ART (see appendix p7-8).

### Counterfactual scenarios

In counterfactual scenarios, we investigate the impact of interventions such as those proposed in the RESOLVE trial^24^ on transmitted and acquired dolutegravir resistance. Specifically, we compared our baseline scenario, where current treatment guidelines in South Africa are modelled, with two alternative scenarios, where measures to reduce dolutegravir resistance are modelled: i) upon entering failing dolutegravir compartments, people are switched to initiating PI-based ART within an average time of 6 or 12 months, and ii) upon an average time of 6 or 12 months on failing dolutegravir-based ART, people with intermediate or high level dolutegravir resistance are switched to PI-based ART within an average time of 6 months (representing the duration taken for druglevel- and resistance testing, receiving results, and implementing treatment adjustments). These two modelled scenarios correspond closely to the RESOLVE interventions in which i) upon detection of viral failure on dolutegravir-based ART, all people are immediately switched to PI-based ART, and ii) upon detection of viral failure on dolutegravir-based ART, all people undergo drug-level- and resistance testing, followed by switch to PI-based ART in case of resistance, and continue on dolutegravir-based ART otherwise. The appendix (p15) provides further details on the scenarios.

### Sensitivity analyses

We investigated the effects of mutation acquisition and reversion rates by varying viremia duration on dolutegravir-based ART rates from 2 to 6 months and the time for dolutegravir DRMs to revert to wild type from 6 months to 20 years. Further, we varied the hazard ratio for acquiring dolutegravir DRMs, comparing NRTI susceptible with resistant, from 1 (no impact) to 10. Similarly, we varied the hazard ratio for failing dolutegravir-based ART associated with high-level resistance from 2 to 4. We varied the probability of transmitting resistant strains from 100% (as likely as susceptible strain transmission) to 5% (20 times less likely than susceptible strain transmission). Finally, we varied the proportion of PWH with detectable drugs failing on dolutegravir-based ART from 0.3 to 1.

In addition to varying parameters one by one, we performed a multidimensional sensitivity analysis by implementing a Monte Carlo estimation of the first-order and total Sobol indices,^25^ which are quantitative measures to assess the importance of parameters (and their interactions in the case of total Sobol indices) on the variability of modelled outcomes. The indices help determine relative parameter importance and identify knowledge gaps (appendix p14, p16).

## Results

The model predicts a steep increase in the number of individuals on dolutegravir-based ART after its introduction in 2020, followed by a more modest but steady increase with numbers approaching eight million PWH on dolutegravir by 2040. The proportion with viral suppression in PWH on dolutegravir-based ART is estimated at around 93%, with those virally non-suppressed (viral load > 1’000 copies per ml) comprised of an increasing proportion with dolutegravir resistance (Figure 2). Among those on dolutegravir-based ART, CD4 levels continue to improve (appendix, p 14)

**Figure 2:**
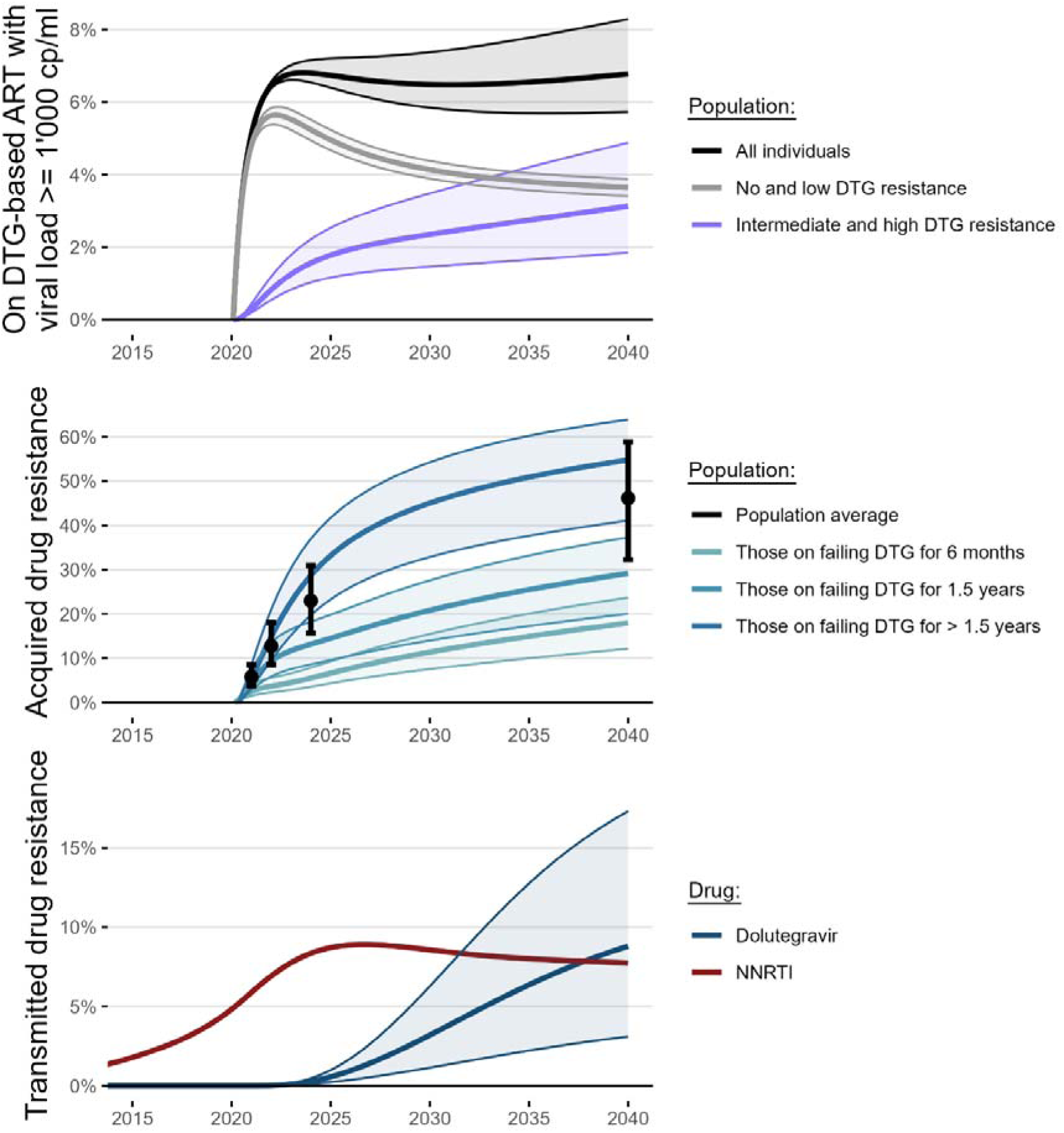
Modelled viral suppression on DTG-based ART, acquired and transmitted dolutegravir resistance. . Solid lines show baseline parameterisation regarding mutation reversion rates, impact of NRTI resistance on dolutegravir DRM acquisition, impact of dolutegravir resistance on its efficacy, transmission probability of dolutegravir DRMs in an HIV transmission event, and proportion with detectable druglevelsdrug levels on failing dolutegravir-based ART. Shaded areas correspond uncertainty intervals (see appendix p12). Dolutegravir rollout was started in 2020. Viral suppression (viral load below 1’000 copies per ml) on dolutegravir-based ART is high; depending on the model assumptions for dolutegravir resistance (see uncertainty range), between 6% and 8% of people on dolutegravir-based ART may be virally unsuppressed. Acquired dolutegravir resistance is defined as proportion of people on a failing dolutegravir-based ART with intermediate- or high-level dolutegravir resistance. Transmitted dolutegravir resistance is defined as proportion of newly diagnosed people with intermediate- or high-level dolutegravir resistance.

### Acquired and transmitted dolutegravir resistance

The model predicts that acquired dolutegravir resistance in people with virologic failure on dolutegravir-based ART will increase rapidly, from 18.5% (uncertainty range 12.5% to 25.4%) in 2023 to 46.2% (uncertainty range 32.9% to 58.9%) in 2040 (Figure 2). There were substantial differences in the predicted levels of acquired dolutegravir resistance depending on the duration of failing dolutegravir-based ART, with a lower prevalence in people experiencing virologic failure for 6 months or less and a higher prevalence among those experiencing failure for over 1.5 years. For transmitted drug resistance, the model predicts a slow but steady increase from 0.1% (uncertainty range 0.1% to 0.2%) in 2023 to 8.8% (uncertainty range 4.4% to 17.3%) in 2040. The rise of transmitted NNRTI resistance will be broken by introducing dolutegravir, but transmitted NNRTI resistance will remain prevalent at 7.7% in 2040 (Figure 2)

### Counterfactual scenarios

We find that reducing the duration PWH remain viraemic whilst on dolutegravir-based ART would substantially reduce acquired dolutegravir resistance at the population level. Immediately switching to PI-based ART upon detecting of viral failure may reduce acquired dolutegravir resistance levels by 2040 from 46.2% to 14.6% and 8.2%, depending on time to detection of viral failure. If using the mitigation strategy with GRT informed ART adjustment upon detection of viral failure, projected acquired dolutegravir resistance may be reduced to 11.3% and 6.3% (Figure 3). Transmitted dolutegravir resistance may be more than halved by 2040, from 8.8% to 2.1% and 1.1% in case of immediate switching to PI-based ART, or to 3.0% and 2.3% in case of GRT informed ART adjustment (Figure 3). Further details are available from the appendix (p15).

**Figure 3:**
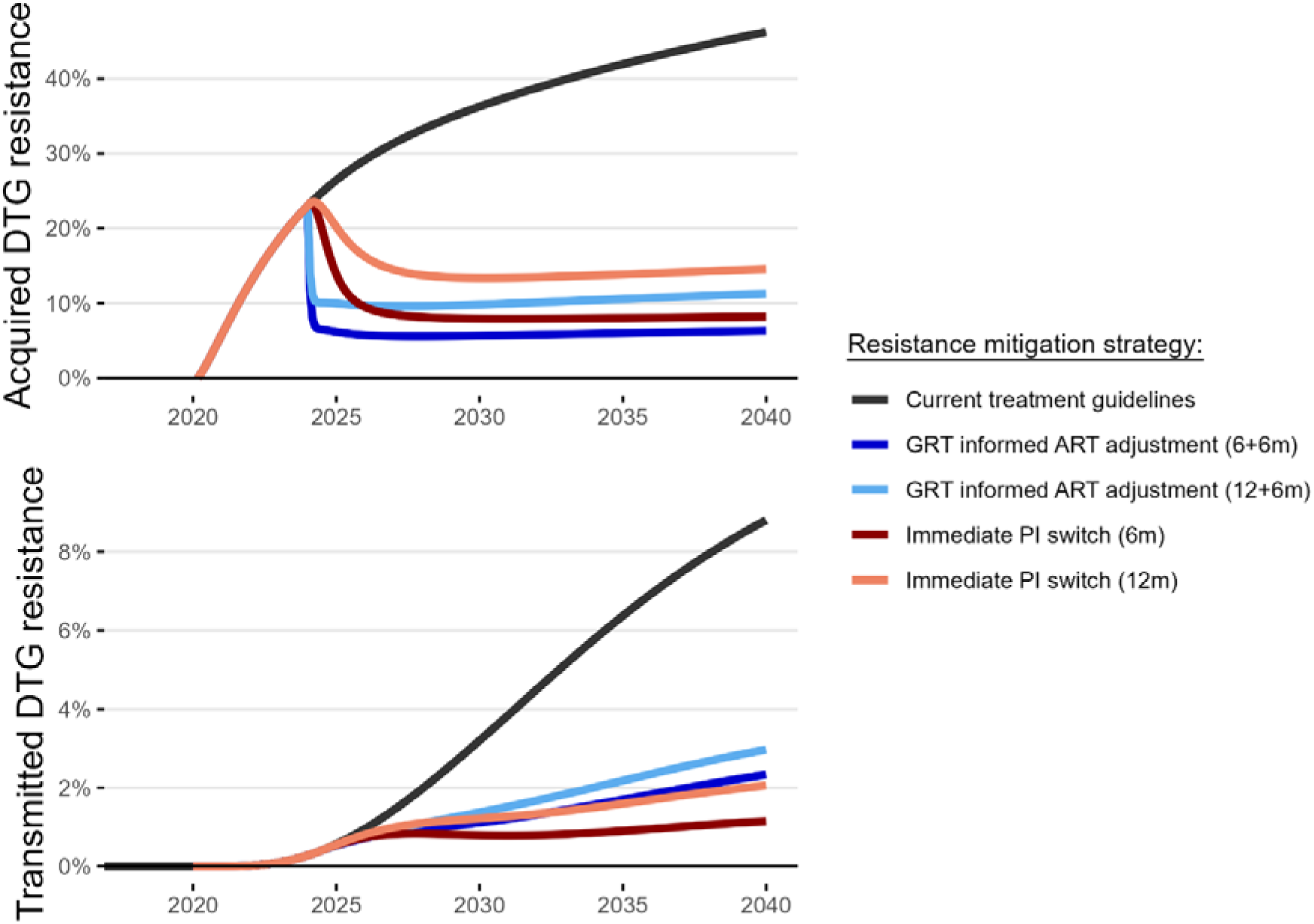
Counterfactual scenario, modelling resistance emergence mitigation strategies. Both strategies reduce the duration with viremia on dolutegravir-based ART. GRT-informed ART adjustment includes druglevel testing and genotypic resistance testing upon detecting virologic failure, followed by switching to PI-based ART in case of intermediate or high level dolutegravir resistance. Immediate PI switch includes switching people with virologic failure on dolutegravir-based ART immediately to PI-based ART. Modelled scenarios include projections assuming 6 and 12 months until virologic failure is detected (6 months: dark blue and dark red lines; 12 months: light blue and light red lines) upon which the respective strategy is implemented immediately. In case of drugleveldrug level-and genotypic resistance testing, we assume an additional average duration of 6 months from detection of virologic failure to informed treatment adjustment. Acquired dolutegravir resistance is defined as proportion of people on a failing dolutegravir-based ART with intermediate- or high-level dolutegravir resistance. Transmitted dolutegravir resistance is defined as proportion of newly diagnosed people with intermediate- or high-level dolutegravir resistance..

### Sensitivity analyses

We assessed the impact of key model parameters one by one and also performed a multivariate sensitivity analysis. Perturbing one parameter at a time showed that dolutegravir resistance outcomes are affected by several variables, but the results from the main analysis were robust.

However, substantial deviations were observed for extreme parameter values, for example, assuming that the probability of transmission of dolutegravir resistance mutations was twenty times less likely than for NNRTIs, assuming much higher acquisition rates of dolutegravir resistance mutations or higher or lower reversion rates or proportion of people with detectable drug levels than in the main analysis. (appendix p16-18)

The variance-based global sensitivity analysis showed that predicted dolutegravir ADR was strongly influenced by assumptions regarding the proportion of people with detectable drug levels (Total Sobol’ sensitivity index 0.34; 95% CI 0.30 to 0.38), and mutation acquisition rates (0.30; 95% CI 0.27 to 0.33). Assumptions for the impact of NRTI resistance on dolutegravir resistance acquisition and of dolutegravir resistance on efficacy also affected ADR predictions (0.19; 95% CI 0.17 to 0.21 and 0.16; 95% CI 0.14 to 0.18, respectively). The assumed dolutegravir DRM transmission probability compared to NNRTIs was the most important factor for the uncertainty in predicted dolutegravir TDR (0.64; 95% CI 0.56 to 0.70). Mutation reversion rates also affected TDR predictions (0.20; 95% CI 0.16 to 0.23).

## Discussion

### Summary of findings

The transition to dolutegravir-based antiretroviral therapy (ART) has contained NNRTI resistance as a threat to the long-term effectiveness of ART. Still, reliance on INSTIs for first-line and second-line ART and prevention of HIV transmission carries the risk of emerging drug resistance to dolutegravir. We updated MARISA, a mathematical model of antiretroviral drug resistance, to study the emergence of resistance to dolutegravir in South Africa.^16,17^ The model fitted the South African data well, with the modelled proportion of PWH with viral suppression (around 93%) in line with empirical data^26^ and the 92% estimate reported by UNAIDS.^27^ We found that the prevalence of acquired and transmitted dolutegravir resistance will likely increase. The 10% threshold of transmitted drug resistance, above which WHO recommends the replacement of a drug, could be reached by 2035, depending on the monitoring and switching strategies in place. During the same period, the prevalence of transmitted NNRTI will decline and then plateau at around 8%. Timely switching of PWH experiencing virologic failure to PI-based ART would substantially reduce acquired and transmitted drug resistance.

### Modelled resistance in context with empirical data

Currently, viral suppression rates on dolutegravir are high. Although acquired drug resistance is rare, it can occur, especially in individuals with a compromised NRTI backbone.^6,7^ Transmission of dolutegravir resistance has also recently been documented.^23^ Programmatic data on dolutegravir resistance in resource-limited settings are scarce. Nationally representative HIV drug resistance surveys based on remnant routine diagnostic viral load samples in South Africa suggest increasing levels of dolutegravir resistance. Steegen et al.^9^ found that in samples with viral load >1000 copies/mL and detectable dolutegravir, resistance to dolutegravir increased from 2.7% in 2021 to 11.1% in 2022, consistent with our predictions. Similar proportions of PWH with resistance mutations on failing dolutegravir-based ART have been recently reported by Tschumi et al. in Lesotho,^7^ and by the WHO from other countries in Southern Africa.^10^

### Duration on failing regimen

Our model predicts that acquired resistance may increase relatively quickly in the coming years. However, the predicted increase in resistance strongly depends on how long people with HIV remain viraemic on a failing dolutegravir-based regimen. There are often delays in switching, and some PWH may not be switched at all. A study linking medical and laboratory data from PWH on first-line ART in 52 South African clinics from 2007 to 2018 reported that only about 40% of PWH with confirmed virological failure were switched.^28^ Among those who switched, the median time to switch was about 16 months, with an interquartile range of 8 to 29 months.^28^ Similarly, an analysis of a South African private sector ART program found that the median time from virologic failure to switching to second-line ART was about 13 months (interquartile range 8 to 22 months), with delays in switching associated with increased mortality.^29^ Other studies from sub-Saharan Africa, including data from Central Africa, East Africa and West Africa, also found sub-optimal switching in adult HIV cohorts after virologic failure, with delays typically ranging from 4 to 17 months.^30–33^

Real-world evidence on effective switching strategies will come from the ongoing RESOLVE trial^24^ in public-sector HIV clinics in Uganda and South Africa. The trial compares universal switching to PI-based second-line ART with switching guided by genotypic resistance tests, urine tenofovir adherence assays, and standard of care. In line with the association between the time spent on a failing regimen and resistance development, the model shows that such resistance mitigation strategies involving rapid switching to PI-based ART could effectively curb the increase in acquired and slow down the increase in transmitted dolutegravir resistance. Both strategies could keep the prevalence of dolutegravir resistance below 10% throughout the modelled period, i.e. up to 2040, in case of switching or drugleveldrug level- and resistance testing, respectively, within 6 months after virologic failure. Of note, the total number of people on PI-based ART differs strongly between the two mitigation strategies. Implementing the immediate PI-switch strategy would result in almost half of all people on ART in South Africa on PI-based ART by 2040.

### Strengths and limitations

Strengths of our study include the combination of modelling the population-level epidemiology of the HIV epidemic in South Africa and of INSTI, NNRTI and NRTI drug resistance, taking into account the differences in the genetic barrier of these drug classes, and the model parametrisation using data from a large collaborative study of dolutegravir resistance.^6^ Further, the model allowed us to assess the impact of public health interventions in counterfactual scenarios. Limitations include large parameter uncertainty, particularly regarding mutation pathways and acquisition- and reversion rates, which we addressed in the sensitivity analyses. The global sensitivity analysis helped unravel the relative importance of these parameters, and identified key knowledge gaps in our understanding of dolutegravir resistance dynamics. The limited data available on drug resistance mutation and their accumulation is a further limitation that may only be addressed in the future when more data becomes available. There is also uncertainty about the interactions between DRM’s impact on viral fitness, including the role of compensatory mutations and the transmission of DRMs. This uncertainty affects transmitted drug resistance more than acquired, as reflected in the broader range of modelled outcomes for transmitted drug resistance. The assumed treatment cascade is a simplification. For example, more complicated treatment histories or the dis- and re-engagement in care^34^ that may favour the emergence of resistance were not explicitly modelled.

### Conclusions

In conclusion, this modelling study indicates that dolutegravir resistance may increase considerably in South Africa: without changes in the management of virologic failure on dolutegravir-based ART, the emergence of transmitted dolutegravir resistance appears to be a question of when, not if, and acquired dolutegravir resistance may increase rapidly in the near future. Drug resistance mitigation strategies for those with virologic failure on dolutegravir-based ART, such as GRT-informed ART adjustment, could strongly reduce acquired and transmitted dolutegravir resistance if virologic failure is detected rapidly. We conclude that dolutegravir resistance surveillance including enhanced viral load monitoring and genotypic resistance testing should be strengthened, especially in settings with programmatic use of dolutegravir-based ART where people remain longer on failing ART. Greater access to genotypic resistance testing^35^ as well as low-cost rapid point-of-care tests of antiretroviral drug levels,^36^ and making generic darunavir/ritonavir available for around 200 US dollars per patient per year^37^ should increase rates of switching to second-line ART and reduce delays, and thereby curbing the emergence and spread of dolutegravir resistance.

## Data Availability

All data produced in the present work are contained in the manuscript or the appendix.

## Authors’ contributions

RDK, RL, ME, and HFG conceptualised the study. TL, JJ, and RDK curated the data. TL, AH, JJ, and RDK devised the methodology. TL, NH, and JJ conducted the formal analysis and validation. TL, NH, and JJ managed the data. RDK provided project administration. AH and ME contributed resources. TL, NH, and JJ applied and adapted the software. HFG, RL, ME, LFJ, AH, and RDK provided supervision. TL created the figures. TL and RDK wrote the original draft of the manuscript. All authors reviewed the manuscript. TL, NH and RDK have directly accessed and verified the underlying data reported in the manuscript. All authors had the final responsibility for the decision to submit for publication.

## Acknowledgements

This work was supported by the University Research Priority Program ‘Evolution in Action’, the Swiss National Science Foundation (324730_207957), and the NIH National Institute of Allergy and Infectious Diseases under award number R01AI152772. We thank the DTG RESIST study group for helpful discussions.

