## Appendix for "Predicting emergent Dolutegravir resistance in South Africa: A modelling study"

### 1 The MARISA model

#### 1.1 MARISA model

MARISA (Modelling Antiretroviral drug Resistance In South Africa) is a mechanistic, compartmental model developed to capture the dynamics of HIV NNRTI resistance among adults in South Africa [1], and is described in detail elsewhere [1, 2]. It models the continuum of care, the disease progression, acquisition and reversion of NRTI and NNRTI resistance, and their impact on the efficacy of the ART regimens. The model was calibrated using different sources of data: 1) cohort data about more than 54,000 people living with HIV from IeDEA collaboration [3, 4], 2) data from literature, and 3) general HIV estimates at the country scale produced by the Thembisa model. The Thembisa model is a compartmental model providing UNAIDS with estimates on the South African HIV epidemic [5].
The model was adapted to investigate the impact of the introduction of DTG-based regimens in South Africa in 2020[2]. The changes included incorporating DTG-based regimen into the continuum of care. The adapted MARISA model is split in 4 dimensions: 1) care stages (15 levels), 2) disease progression, characterised by the CD4 counts (4 levels), 3) gender (2 levels), 4) NRTI and NNRTI resistance.

#### 1.2 MARISA model for DTG resistance

The model was further adapted to investigate complex resistance dynamics. The binary resistance dimension was hereby extended to a "resistance genotype", represented as key drug resistance mutations against INSTIs, while also retaining NRTI and NNRTI resistance. Furthermore, we subdivided the "Failed" compartment on DTG-based ART into "Failed recent", "Failed intermediate", and "Failed long" (see section 2.2.1), extended the cascade of care dimension by adding an "Out of care" stage (see section 2.2.2), and take into account the proportion of people on treatment with undetectable drug levels (see table 6).
In short, the first dimension of the model accounts for the continuum of care, starting at HIV-infection of susceptible individuals and diagnosis, followed by first (NNRTI- or DTG-based) and second (PI-based) line treatment. Each regimen has three stages; treatment initiation ("Treated") with subsequent virological suppression ("Suppressed") or failure ("Failed"), whereby "Failed" is composed of three compartments reflecting time on failing treatment (see section 2.2.1). Before 2020, all individuals receive a NNRTI-base first-line regimen and switch to the second-line PI-based regimen in case of confirmed virological failure. From 2020, the DTG-based regimen is used as a first-line regimen for all populations. All individuals who are on NNRTI-based regimen can transition to DTG-based regimen (figure 1). The second dimension splits individuals in 4 classes according to CD4 counts: 1) $CD4 > 500$ cells/$\mu L$, 2) $350 < CD4 < 500$ cells/$\mu L$, 3) $200 < CD4 < 350$ cells/$\mu L$ and 4) $CD4 < 200$ cells/$\mu L$. The third dimension makes the distinction between male and female. The fourth dimension comprises the different resistance mutations, collectively making up the resistance genotype. Its structure and parametrization are described in section 2.4.
We used the following indices to indicate a layer of a dimension: $j$ for the second dimension ($j = 1, 2, 3, 4$), $k$ for the third dimension ($k = 1, 2$) and $l$ for the fourth dimension ($l = 1, ..., x$).

### 2 Parameters and rates of the DTG MARISA model

Parameters and rates regarding the continuum of care and disease progression dimensions have been described in detail before[1, 2].

#### 2.1 Diagnosis, treatment initiation and switching rates to PI-based regimen

Diagnosis rates depend on gender and CD4 classes and treatment initiation rates depend on CD4 classes. They have been described in detail in [1]. In short, diagnosis rates are constant from 2016, while the treatment initiation rate has been adapted in order to model the impact of the Treat-All policy. Treatment initiation rates are increased for the first three first CD4 count classes from 2017 to 2022 in order to have identical rates irrespective of CD4 counts from 2022. Switching rates from unsuppressed first line regimen (NNRTI-based and/or DTG-based) to PI-based regimen $\gamma^k_{F_{NNRTI} \to T_{PI}}$ were rescaled in order to reflect PI-coverage in South Africa ($\sim$ 4% in 2016 according to [6]). Treatment initiation rates are described in detail in [2]. In short, treatment initiation rates on DTG after 2020 are considered equal to the treatment initiation rates on NNRTI prior to 2020. As a consequence of the Treat-All policy implemented in 2017, treatment initiation rates of HIV infected individuals increase until 2022, after which they remain constant (see [2] supplemental information S1 Text, Section 2.2). Switching rates from NNRTI- to DTG-based regimens were fixed for both switching from

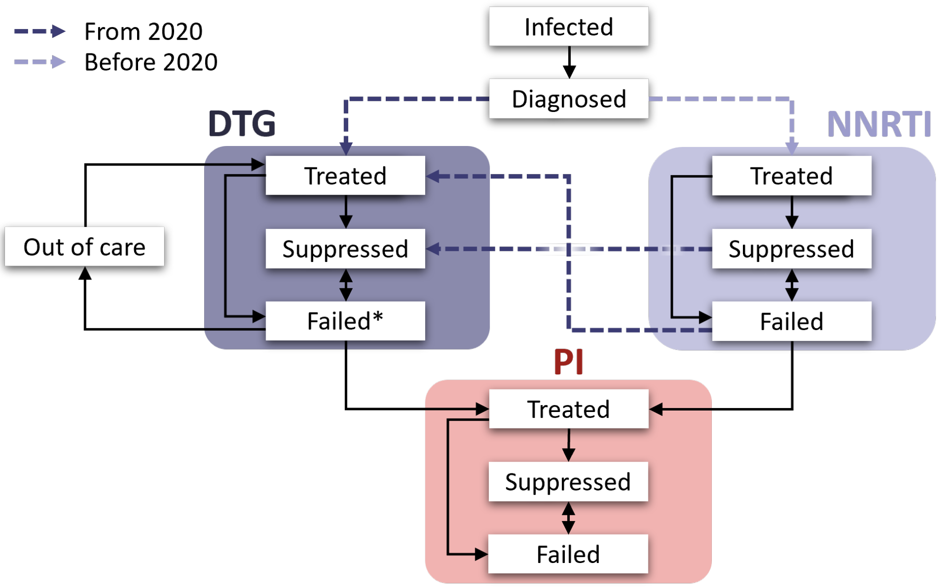

**Figure 1:** Cascade of care dimension in DTG MARISA. On DTG-based ART, the "Failed" compartment is further subdivided into "Failed recent", "Failed intermediate", and "Failed long".

suppressed or failing NNRTI-based regimen to 1 year$^{-1}$. Individuals switching from suppressed NNRTI-regimen would remain suppressed, while individuals switching from failing NNRTI-based regimen would start DTG in the "Treated" compartment (see figure 1).

#### 2.2 Rates related to continuum of care and disease progression

Rates related to continuum of care and disease progression have been described and published before[1, 2]. In short, rates related to disease progression $\nu_{CD4}$ and $\tilde{\nu}_{CD4}$ as well as rates related to continuum of care $\gamma$, which respectively model transition from one to another CD4 class and transition from one to another care stage, were estimated using observational cohort data from IeDEA-SA collaboration. Mean estimates and 95% confidence intervals (95%CI) are reported in Table 1 and 2.

**Table 1:** Rates related to disease progression. Rates are in month$^{-1}$. CD4 classes correspond to: 1: $CD4 > 500$ cells/$\mu L$; 2: $350 < CD4 < 500$ cells/$\mu L$; 3: $200 < CD4 < 350$ cells/$\mu L$; 4: $CD4 < 200$ cells/$\mu L$.

| Parameter | Description | Values [95% CI] | | |
|---|---|---|---|---|
| *Parameters related to disease progression* | | | | |
| | | CD4 class | | |
| | | $1 \rightarrow 2$ | $2 \rightarrow 3$ | $3 \rightarrow 4$ |
| $1/\nu^{I}_{CD4}$ | Average time to progress from one to another CD4 class, at $I$ (taken from [7]) | 60 | 36 | 42 |
| $1/\nu^{D}_{CD4}$ | Average time to progress from one to another CD4 class, at $D$ (taken from [7]) | 60 | 36 | 42 |
| $1/\nu^{T_{NNRTI}}_{CD4}$ | Average time to progress from one to another CD4 class, at $T_{NNRTI}$ | 47 [42,54] | 30 [28,34] | 60 [55,66] |
| $1/\nu^{F_{NNRTI}}_{CD4}$ | Average time to progress from one to another CD4 class, at $F_{NNRTI}$ | 18 [16,20] | 15 [14,16] | 22 [21,24] |
| $1/\nu^{T_{PI}}_{CD4}$ | Average time to progress from one to another CD4 class, at $T_{PI}$ | 32 [14,72] | 22 [12,43] | 33 [17,64] |
| $1/\nu^{F_{PI}}_{CD4}$ | Average time to progress from one to another CD4 class, at $F_{PI}$ | 14 [8,26] | 15 [8,27] | 16 [10,25] |
| | | $1 \leftarrow 2$ | $2 \leftarrow 3$ | $3 \leftarrow 4$ |
| $1/\tilde{\nu}^{T_{NNRTI}}_{CD4}$ | Average time to progress from one to another CD4 class, at $T_{NNRTI}$ | 16 [15,17] | 16 [15,17] | 18 [17,19] |
| $1/\tilde{\nu}^{S_{NNRTI}}_{CD4}$ | Average time to progress from one to another CD4 class, at $S_{NNRTI}$ | 17 [16,17] | 14 [14,14] | 9 [9,10] |
| $1/\tilde{\nu}^{T_{PI}}_{CD4}$ | Average time to progress from one to another CD4 class, at $T_{PI}$ | 16 [9,27] | 19 [11,31] | 41 [23,73] |
| $1/\tilde{\nu}^{S_{PI}}_{CD4}$ | Average time to progress from one to another CD4 class, at $S_{PI}$ | 17 [13,21] | 14 [11,17] | 7 [6,10] |

##### 2.2.1 Time on failing DTG-based ART

In order to model time on failing DTG-based ART, the "DTG Failed" compartment was split into the three compartments "DTG Failed recent" ($F_{DTG_{rec}}$), "DTG Failed intermediate" ($F_{DTG_{int}}$), and "DTG Failed long" ($F_{DTG_{long}}$), which are identical regarding re-suppression rates, out of care rates, mortality, and CD4 progression. Progression from $F_{DTG_{rec}}$ to $F_{DTG_{int}}$ ($\gamma_{F_{DTG_{rec}} \rightarrow F_{DTG_{int}}}$) is 1/6, i.e., a time of on average 6 months for those who did not either progress to PI-based ART, re-suppressed on DTG-based ART, or died. Similarly, progression $\gamma_{F_{DTG_{int}} \rightarrow F_{DTG_{long}}}$ is 1/12, which corresponds to taking on average 12 months for those not advancing to other compartments.

##### 2.2.2 Out of care

Out of care dynamics were implemented starting with DTG-rollout to adjust for the impact of this sub population of people with viraemia, but without selective pressure imposed on the virus by the ART regimen on both acquired and transmitted drug resistance. Hereby, people on failing DTG-based ART may drop out of care with a rate of 109 per 1000 person-years[8], and the average time in the "Out of care" compartment before re-entering the "DTG Treated" compartment is 22.8 months[9] (see table 6).
Disease progression and mortality in "Out of care" are assumed to be equal to those in "DTG Failed" compartments (see $1/\nu^{F_{NNRTI}}_{CD4}$, table 1, and $\tilde{\mu}^{i}_{F_{NNRTI}/F_{PI}/O}$, table 6). People in "Out of Care" are considered as infectious, not acquiring any drug resistance, and allowing reversion (see section 2.3.6).

**Table 2:** Rates related to transition between care stages. Rates are in month$^{-1}$. CD4 classes correspond to: 1: $CD4 > 500$ cells/$\mu L$; 2: $350 < CD4 < 500$ cells/$\mu L$; 3: $200 < CD4 < 350$ cells/$\mu L$; 4: $CD4 < 200$ cells/$\mu L$.

| Parameter | Description | Values [95% CI] | | | |
|---|---|---|---|---|---|
| *Parameters related to care stages* | | | | | |
| | | CD4 class | | | |
| | | 1 | 2 | 3 | 4 |
| $1/\gamma_{T_{NNRTI} \to S_{NNRTI}}$ | Time from $T_{NNRTI}$ to $S_{NNRTI}$ | 3.4 [3.3,3.6] | 3.5 [3.3,3.7] | 3.6 [3.4,3.7] | 3.9 [3.8,4.1] |
| $1/\gamma_{T_{NNRTI} \to F_{NNRTI}}$ | Time from $T_{NNRTI}$ to $F_{NNRTI}$ | 23.4 [20.1,27.3] | 22.8 [19.6,26.4] | 18.9 [17.5,20.4] | 12.9 [12.3,13.5] |
| $1/\gamma_{S_{NNRTI} \to F_{NNRTI}}$ | Time from $S_{NNRTI}$ to $F_{NNRTI}$ | 176.3 [157,197.9] | 133.8 [118.6,150.8] | 62.1 [57,67.6] | 22.1 [20.2,23.9] |
| $1/\gamma_{F_{NNRTI} \to S_{NNRTI}}$ | Time from $F_{NNRTI}$ to $S_{NNRTI}$ | 6.4 [5.5,7.4] | 12.9 [11,14.9] | 14.3 [12.9,15.9] | 18.2 [16.3,20.2] |
| $1/\gamma_{F_{NNRTI} \to T_{PI}}$* | Time from $F_{NNRTI}$ to $T_{PI}$ | 467.5 [243,898.9] | 376 [240.4,589.9] | 258.9 [200.7,334.6] | 166.4 [140,199] |
| $1/\gamma_{T_{PI} \to S_{PI}}$ | Time from $T_{PI}$ to $S_{PI}$ | 3.8 [2.7,5.2] | 3.8 [2.6,5.5] | 4 [3,5.3] | 5 [4,6.4] |
| $1/\gamma_{T_{PI} \to F_{PI}}$ | Time from $T_{PI}$ to $F_{PI}$ | 14.3 [7.8,26.8] | 14 [7.3,27] | 11.8 [7.8,18] | 7.6 [5.9,9.9] |
| $1/\gamma_{S_{PI} \to F_{PI}}$ | Time from $S_{PI}$ to $F_{PI}$ | 61.4 [30.8,122.8] | 40.9 [21.4,78.9] | 40 [21.4,74.3] | 19.1 [9,40] |
| $1/\gamma_{F_{PI} \to S_{PI}}$ | Time from $F_{PI}$ to $S_{PI}$ | 2.3 [1.1,4.1] | 12.9 [3.2,51.3] | 5.5 [2.8,11.3] | 11.7 [4.8,28] |
| *Parameters related to time on failing DTG-based ART* | | | | | |
| $1/\gamma_{F_{recent} \to F_{int}}$** | Time from recently failing to intermediate time on failing DTG-based ART | 6 | | | |
| $1/\gamma_{F_{int} \to F_{long}}$** | Time from intermediate to long time on failing DTG-based ART | 12 | | | |

Note:

* Switching rates $\gamma^k_{F_{NNRTI} \to T_{PI}}$ are rescaled to reflect PI-coverage in South Africa ($\sim 4\%$ in 2016 according to [6]).

** Time on failing treatment is only considered for DTG-based ART. F1 thus corresponds to DTG only, and starting after the DTG-rollout in 2020.

#### 2.3 Resistance rates

##### 2.3.1 DTG drug resistance mutations (DRMs)

The resistance dimension is adapted to include complex mutational patterns and acquisition pathways. Resistance to NRTI and NNRTI is treated as before[1, 2], and is considered in the resistance genotype. For DTG resistance, the following key mutations were included in the model: G118R, E138K, G140ACS, Q148HKNR, N155H, and R263K. Key mutations for DTG resistance were identified based results from the DTG RESIST study in 599 people on failing DTG-based ART[10], and a rapid scoping review for DTG resistance in 2023[11]. The drug resistance dimension is based on combinations of the "resistance genotype" with the following composition (whereby NNRTI and NRTI positions are populated using previously found rates):

$$G118R - E138K - G140ACS - Q148HKNR - N155H - R263K - NNRTI - NRTI$$

Note that the order of the mutations above does not hold any meaning. Allowed combinations and acquisition patterns are described in the next section.

##### 2.3.2 DTG DRM accumulation pathway

The dimension could thus hold $2^8 = 256$ possible resistance genotypes; however, mutations do not occur randomly. The DTG resistance mutations we allow in the model are derived from the observed cases of DTG resistance in [10]. Figure 2 shows the modelled 12 DTG DRM combinations their accumulation. Including resistance/sensitivity to NRTIs and NNRTIs (see section 2.3.4) results in a total of 48 allowed resistance genotypes, which corresponds to the number of modelled resistance compartments.

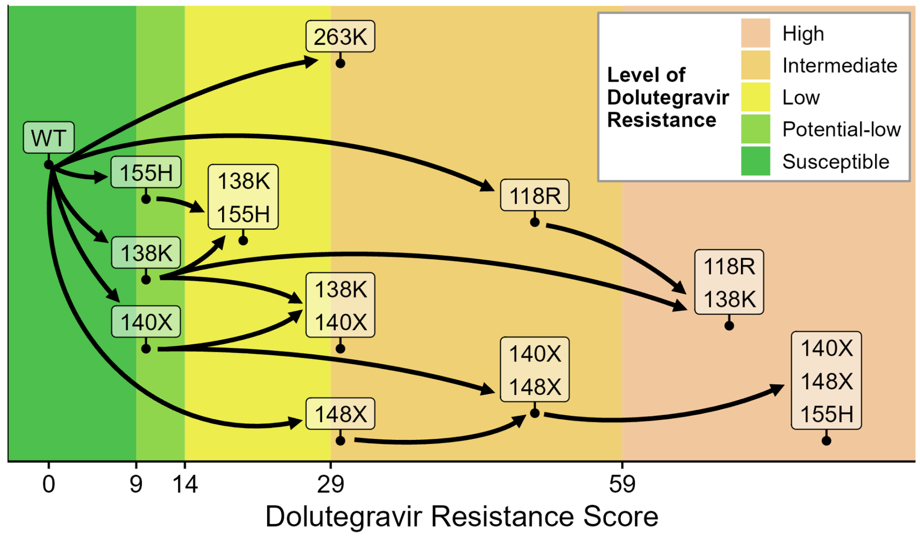

**Figure 2:** DTG resistance mutations included in the DTG MARISA model, including allowed combinations and accumulation pathways. 140X comprises mutations G140G/C/S, 148X comprises G148H/K/N/R. Drug resistance scores and levels are derived from the Stanford resistance algorithm, Version 9.5.1

##### 2.3.3 DTG DRM acquisition rates

Mutation acquisition rates $r_{DRM_m}$, whereby $m$ represents a specific dolutegravir resistance mutation, are derived from the 599 people on failing DTG-based ART in the DTG RESIST study according to equation 1, where $\dot{M}_q(t)$ is the number of observed cases per DTG resistance mutation combination $q$. The mutation combination from which $q$ originates by acquisition of a single mutation is denoted as $q'$, and $U_q$ is the set of all possible $q'$. We use $WT$ (wildtype) for the special case where $q$ is 00000000, i.e., does not have any DTG DRMs, NNRTI, or NRTI resistance.

$$\delta_x = \begin{cases} 1 & \text{if x = WT,} \\ r_{acc} & \text{if x} \neq \text{WT.} \end{cases}$$

$$\dot{M}_q(t) = \sum_{q' \in U_q} M_{q'}(t) * r_{DRM_{q' \neq q}} * \delta_{q'} \tag{1}$$

We include $r_{acc}$ as additional parameter, which represents the observed increased mutation acquisition rate after having acquired at least one resistance mutation. In the DTG RESIST study population, a median of three viral load tests were performed per year, and resistance testing was performed at detection of virologic failure. However, the actual time on failing DTG-based ART is unknown. In our default model we assume the mutations to be observed after 3 months on a failing regimen. Mutation rates depending on different assumptions regarding time on failing DTG-based ART can be found in Table 3. The impact of the assumed time on failing DTG-based ART on model outcomes is further explored in the section 3.4.

**Table 3:** Rates related to drug resistance mutation acquisition. Mutation acquisition rates are increased by $r_{acc}$ upon acquiring at least one mutation. Rates are in month$^{-1}$.

| Parameter | Description | Values | | |
|---|---|---|---|---|
| | | Assumed time on failing DTG-based ART (see eq. 1)* | | |
| | | 2 months | 3 months | 6 months |
| *Mutation acquisition rates* | | | | |
| $r_{G118R}$ | Time to acquire G118R | 0.0014 | 0.0011 | 0.0005 |
| $r_{E138K}$ | Time to acquire E138K | 0.0012 | 0.0010 | 0.0004 |
| $r_{G140ACS}$ | Time to acquire G140ACS | 0.0062 | 0.0047 | 0.0021 |
| $r_{Q148HKNR}$ | Time to acquire Q148HKNR | 0.0040 | 0.0029 | 0.0013 |
| $r_{N155H}$ | Time to acquire N155H | 0.0053 | 0.0041 | 0.0018 |
| $r_{R263K}$ | Time to acquire R263K | 0.0096 | 0.0072 | 0.0032 |
| *Mutation rate increase* | | | | |
| $r_{acc}$ | Mutation rate multiplier if $\geq$ one DTG DRM is present | 214.8 | 210.6 | 203.4 |

Note:
* Data from the DTG RESIST cohort collaboration study [10].

Figure 3 shows the distribution of dolutegravir resistance levels over time on failing dolutegravir-based ART (assuming treatment adherence, and using DTG DRM rates assuming 3 months on failing DTG-based ART in DTG RESIST, see 1).

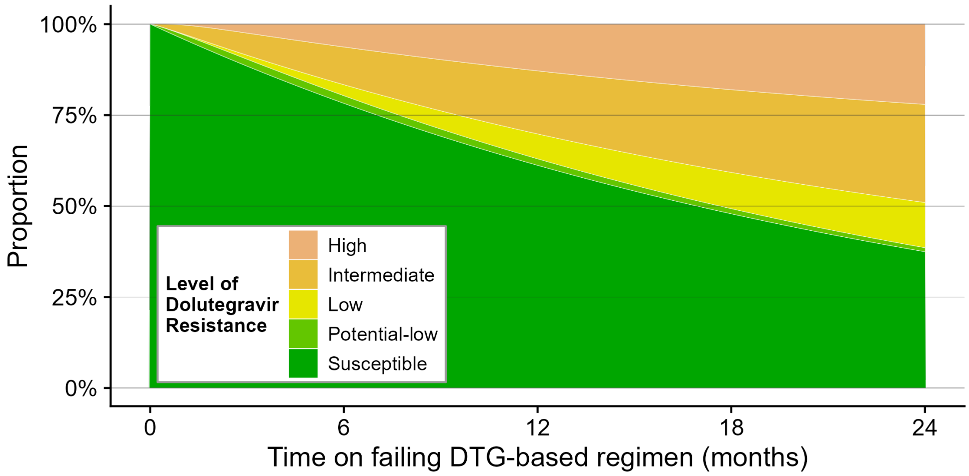

**Figure 3:** Modelled progression in DTG resistance levels over time. Shown is the distribution of dolutegravir resistance levels over time on failing DTG-based ART under the assumption of DTG DRMs accumulating as outlined in 2, and assuming 3 months in equation 1. Drug resistance scores and levels are derived from the Stanford resistance algorithm, Version 9.5.1

##### 2.3.4 NNRTI and NRTI resistance acquisition

The rates for NNRTI and NRTI resistance acquisition have been described in detail before [1, 2]. In short, NRTI-resistance is defined as having both the K65R and the M184V mutations, and was calibrated using results from a meta-analysis that estimates the prevalence of NRTI resistance mutation after 3 years on a failing NNRTI-based first-line regimen [12] (see Table 6). In light of the scarcity of programmatic data of people on failing DTG-based ART, this approach was not feasible for specifically assess NRTI resistance acquisition rates on failing DTG-based ART; we therefore use $\sigma_{res}^{NRTI}$ on DTG-based regimen as well, i.e., we assume NRTI resistance is acquired at the same rate on failing NNRTI-based ART as on failing DTG-based ART.

##### 2.3.5 Resistance acquisition in cascade of care

An individual can acquire NNRTI-resistance when failing a NNRTI-based regimen, and can acquire DTG resistance when failing a DTG-based regimen. NRTI resistance can be acquired on a failing NNRTI- or DTG-based regimen. Acquisition of NRTI and NNRTI resistance is independent of DTG resistance mutations.

##### 2.3.6 Reversion

Reversion to wild-type occurs when virus replication occurs without selective drug pressure, i.e. in the "Infected", "Diagnosed", and "Out of care" compartments, as well as "Failure" for drug resistance positions not part of resistance to the failing regimen (e.g., reversion of NNRTI failing DTG regimen). We assume the same reversion rates for those on failing treatment and those out-of-care.
Rates for NRTI and NNRTI are populated with previously found rates [1, 2] and are listed in Table 6. Reversion rates of DTG resistance mutations are largely unknown, but have been reported to occur[13], and previous work on reversion after treatment interruption following first generation INSTI regimen showed rapid reversion of INSTI DRMs[14]. We assume an average time to reversion of 2 years for all DTG resistance mutations; a wide range is analyzed in the sensitivity analyses, see section 3.4. Reversion along distinct mutational pathways has been shown for NRTI DRMs [15]. For simplicity, DTG resistance mutations are assumed to revert alongside the same pathways as acquisition.

##### 2.3.7 Resistance transmission

In view of the low level of NRTI pre-treatment drug resistance (PDR) [16, 17], we assume that NRTI resistance is not transmitted. The probabilities of DTG resistance mutation transmission are unknown. In the default model, we assume DTG resistance mutations to be transmitted like NNRTI resistance. A range of reduced transmission probabilities for DTG DRMs compared to NNRTI resistance are explored in sensitivity analyses, see section 3.4.

#### 2.4 Structure of the Resistance-Genotype matrices

The resistance dimension is implemented using matrices describing the transitions between resistance genotypes. Two matrices $A_{i,j}$ and $R_{j,i}$ describe the movement between the resistance-genotypes. The Impact of the genotypes on a treatment is given by the matrix $E_{j,trt}$. $A_{1,1}$ and $R_{1,1}$ represent the wildtype, having no resistance mutations and therefore being fully susceptible to all treatments. Table 4 describes all matrices included in modelling transitions in the resistance dimension.

**Table 4:** Description of the compartments used in the model.

| Notation | Description | Definition |
|---|---|---|
| *Resistance matrices* | | |
| $A_{b,c}$ | Matrix with rates to acquire the genotype c from genotype b | |
| $R_{s,t}$ | Matrix with rates to revert to genotype t from genotype s | |
| $E_{f,g}$ | Matrix with the impact of genotype f on treatment g (using the previously implemented $\alpha_1$, $\alpha_2$, and $\alpha_3$ parameters, see note) | g=1: NNRTI, first 3 months, g=2: NNRTI, over 3 months, g=3: PI, g=4: DTG |
| *Additional matrices* | | |
| $P_d$ | Subset of $A_{b,c}$ affected by treatment d | d := NNRTI, DTG, PI |
| $T_{u,v}$ | Matrix for transmission of drug resistance (probability of genotype u being transmitted as genotype v) | |

Note: $\alpha_1$ (Impact of NNRTI-resistance on NNRTI-based ART in the first 3 months on ART) is 1.97[18, 19], $\alpha_2$ (Impact of NNRTI resistance on NNRTI-based ART after 3 months on ART) is 3.24[18, 19], and $\alpha_3$ (Impact of NRTI resistance on DTG-based ART) is 1 [20, 21, 22].

The resistance genotype comprises 8 positions (see 2.3.1), for each included DRM, and NNRTI and NRTI resistance; 00000000 thus represents absence of all DRMs, NNRTI, and NRTI resistance.The matrices are structured as follows (note that for better readability, in the conceptual matrices we only note 3 resistance positions):

$$\mathrm{A}_{b,c} = \begin{array}{c} 000 \\ \vdots \\ 111 \end{array} \overset{\begin{array}{ccc} 000 & \cdots & 111 \end{array}}{\begin{pmatrix} 0 & \cdots & r_{DRM}^{000\to111} \\ \vdots & & \vdots \\ 0 & \cdots & 0 \end{pmatrix}}$$

$$\mathrm{R}_{s,t} = \begin{array}{c} 000 \\ \vdots \\ 111 \end{array} \overset{\begin{array}{ccc} 000 & \cdots & 111 \end{array}}{\begin{pmatrix} 0 & \cdots & 0 \\ \vdots & & \vdots \\ r_{DRM}^{111\to000} & \cdots & 0 \end{pmatrix}}$$

$$\mathrm{E}_{f,g} = \begin{array}{c} 000 \\ \vdots \\ 111 \end{array} \overset{\begin{array}{ccc} NNRTI & DTG & PI \end{array}}{\begin{pmatrix} 1 & 1 & 1 \\ \vdots & \vdots & \vdots \\ \alpha_{impact}^{111\to NNRTI} & \alpha_{impact}^{111\to DTG} & \alpha_{impact}^{111\to PI} \end{pmatrix}}$$

$$\mathrm{P}_{d} = \begin{array}{c} 000 \\ \vdots \\ 111 \end{array} \overset{\begin{array}{ccc} 000 & \cdots & 111 \end{array}}{\begin{pmatrix} 0 & \cdots & \phi_{d}^{000\to111} \\ \vdots & & \vdots \\ 0 & \cdots & 0 \end{pmatrix}}$$

$$\mathrm{T}_{u,v} = \begin{array}{c} 000 \\ \vdots \\ 111 \end{array} \overset{\begin{array}{ccc} 000 & \cdots & 111 \end{array}}{\begin{pmatrix} 1 & \cdots & \phi_{transmission}^{000\to111} \\ \vdots & & \vdots \\ \phi_{transmission}^{111\to000} & \cdots & \phi_{transmission}^{111\to111} \end{pmatrix}}$$

Hereby $r_{DRM}^{000\to111}$ represents resistance acquisition rates (see table 3 for DTG mutation rates, and table 6 for NRTI and NNRTI rates). Reversion rates are represented by $r_{DRM}^{111\to000}$ (see section 2.3 and table 6). Impact of a resistance genotype is depicted as $\alpha_{impact}^{111\to treatment}$, see table 4. $\phi_{d}^{000\to111}$ denotes whether a failing regimen results in selective pressure on a genotype, i.e., $\phi_{DTG}^{WT\to NNRTI} = 0$, while $\phi_{DTG}^{WT\to G118K} = 1$. The transmission of a resistance genotype is denoted as $\phi_{transmission}^{000\to111}$, and reflects the probabilities by which a resistance genotype is transmitted.

#### 2.5 Impact of Resistances on treatments

DTG MARISA uses the E matrix for each resistance genotype and treatment to model the impact of resistance on treatment response. The parameters increase the previously estimated rates of failure $\gamma_{T_{NNRTI}\to F_{NNRTI}}$, $\gamma_{S_{NNRTI}\to F_{NNRTI}}$ and decrease the suppression rates $\gamma_{T_{NNRTI}\to S_{NNRTI}}$ and $\gamma_{F_{NNRTI}\to S_{NNRTI}}$ for resistant individuals.

In order to achieve the same suppression levels as estimated from IeDEA-SA cohort data with the modified rates, the scaling parameter $\alpha_g$ is used, which increase the overall suppression rates and decreases the failing rates. $\alpha_{NNRTI}$ was previously estimated as 1.62 based on overall suppression rates for NNRTI-based regimen (88%) in the IeDEA cohort data [2, 3]. The different failing and suppression rates according to CD4 class $j$ and NNRTI-resistance status $l$ are given in Eq 2-5. The rates $\gamma_{T_{NNRTI}\to F_{NNRTI},g}$, $\gamma_{T_{NNRTI}\to S_{NNRTI},g}$, $\gamma_{S_{NNRTI}\to F_{NNRTI},g}$ and $\gamma_{F_{NNRTI}\to S_{NNRTI},g}$ represent the overall suppression and failure rate for $g$-based ART, as estimated with IeDEA cohort data (see Tables 1 and 2). As impact on treatment for DTG cannot be modelled from the IeDEA cohort data, impact on treatment is assumed to be the same as in NNRTI-based ART. Impact of high level DTG-resistance on DTG-based ART is assumed to be the same as for NNRTI resistance on NNRTI-based ART; the impact of intermediate level DTG-resistance on DTG treatment efficacy is assumed be 50% of the impact of high level resistance.

$$\gamma_{T_{NNRTI}\to F_{NNRTI},g}^{jl} = E[l,g] \cdot 1/\alpha_g \cdot \gamma_{T_{NNRTI}\to F_{NNRTI}} \tag{2}$$

$$\gamma_{T_{NNRTI}\to S_{NNRTI},g}^{jl} := 1/3 - \gamma_{T_{NNRTI}\to F_{NNRTI},g}^{j,l} \tag{3}$$

$$\gamma^{jl}_{F_{NNRTI}\to S_{NNRTI},g} = 1/E[l,g] \cdot \alpha_g \cdot \gamma_{F_{NNRTI}\to S_{NNRTI}} \quad (4)$$

$$\gamma^{jl}_{S_{NNRTI}\to F_{NNRTI},g} = E[l,g] \cdot 1/\alpha_g \cdot \gamma_{S_{NNRTI}\to F_{NNRTI}} \quad (5)$$

#### 2.6 DTG-efficacy and impact of NRTI-resistance on DTG

The efficacy of DTG-based regimens has been a subject of extensive study. In the NAMSAL study, after adjusting for CD4 counts and baseline NNRTI resistance, an odds ratio (OR) of 1.02 was found between NNRTI- and DTG-based regimens (assuming no resistance) [23]. We therefore employ equal treatment efficacy in our model for NNRTI- and DTG-based regimens (a range of other efficacies for DTG-based regimen has been previously investigated in MARISA [2]).
The impact of NRTI resistance on the efficacy of DTG-based regimens has been investigated in the NADIA trial, where viral suppression rates for NRTI-resistant individuals starting a DTG-based regimen were comparable to those of NRTI-sensitive individuals [24]. We thus model NRTI resistance having no impact on DTG efficacy.
According to the large DTG RESIST cohort collaboration, NRTI resistance increases the risk of developing DTG resistance in those with viremia, with an adjusted odds ratio of 13.4 for intermediate or high-level NRTI resistance (95% CI 4.55–39.7) [10]. Therefore, we model NRTI resistance to increase the rates of acquiring DTG resistance. In the default model, the presence of NRTI resistance quadruples the hazard for DTG resistance; in the sensitivity analyses, we cover a wide range from NRTI resistance having no impact on DTG resistance acquisition, to ten times higher hazard 3.4.

#### 2.7 Impact of regimen backbone

As a simplifying assumption, all individuals that transition to DTG-based regimen are assumed to have received a NNRTI-drug combined with TDF and 3TC/FTC and to keep this NRTI-backbones combination after transitioning to DTG-based regimen. This assumption is motivated by the expected reluctance of clinicians to prescribe zidovudine (AZT) for TDF-experienced individuals transitioning to DTG. In the case where NRTI backbones are adapted when transitioning to DTG, the model might overestimate the impact of NRTI-resistance on DTG-based regimen. In counterfactual scenarios, we assess the impact of an optimized backbone regimen for people switching from NNRTI- to DTG-based ART, see section 3.3.

#### 2.8 Other parameters: HIV transmission and mortality

Parameters regarding HIV transmission and mortality have been previously described [1, 2]. Briefly, MARISA accounts for sexual HIV transmission both for sex between men and women, and for sex between men. The model includes different transmission risks per intercourse, and assumes higher risk behaviour in undiagnosed individuals. HIV transmission parameters were sourced from the literature ((see Table 6) or estimated using results from the Thembisa model (see Table 6).
Mortality was modeled based on CD4 counts and treatment stage. Relative mortality estimates were previously obtained from the literature (see Table 6), and a scaling parameter for the mortality risk among suppressed individuals with CD4 $>$ 500 copies/ml was fitted to HIV mortality estimates from the Thembisa model. Further details on HIV transmission and mortality can be found elsewhere [1].

**Table 5:** Parameters estimated from outputs of the Thembisa model.

| Parameter | Description | Values |
|---|---|---|
| $\beta_u$ | Number of unprotected sexual acts per month (for undiagnosed individual) | 3.1 |
| $\beta_d$ | Number of unprotected sexual acts per month (for diagnosed individual) | 1.24 |
| $\gamma_{I\to D}(2016)/\gamma_{I\to D}(2005)$ | Ratio of diagnosis rates between 2005 and 2016 | 4.4 |
| $1/\gamma_{I\to D}(2005)$ | Time to diagnosis in 2005 (months) | 26 |
| $1/\gamma_{D\to T_{NNRTI}}(2005)$ | Time to ART initiation in 2005 (months) | 60 |
| $\mu_0$ | Mortality risk (in $(\text{month} \cdot 1000 \text{ people})^{-1}$) for a suppressed individual with $CD4 > 500$ cells/$\mu L$ | 0.08 |

#### 2.9 Parameters from the literature

**Table 6:** Parameters collected from the literature. As mortality estimates in the fourth CD4 class vary according to the proportion of people with $CD4 < 50$ cells/$\mu L$, lower and upper bounds are given (see [1] S1 File Section 1.2 for more details). The mortality risk $\mu_X^j$ in CD4 class $j$ ($i = 1, \ldots, 4$) and care stage $X$ ($X = I, D, T_{NNRTI}, \ldots$) is given by: $\mu_X^j = \mu_0 \cdot \tilde{\mu}_X^j$. CD4 classes correspond to: 1: $CD4 > 500$ cells/$\mu L$; 2: $350 < CD4 < 500$ cells/$\mu L$; 3: $200 < CD4 < 350$ cells/$\mu L$; 4: $CD4 < 200$ cells/$\mu L$.

| Parameter | Description | Values | Ref | | |
|---|---|---|---|---|---|
| *Resistance parameters* | | | | | |
| $1/\sigma_{res}^{NNRTI}$ | Time to acquire NNRTI-resistance (months) | 5 | [25, 26, 27, 28, 29] | | |
| $1/\sigma_{res}^{NRTI}$ | Time to acquire NRTI-resistance (months) | 40 | [12] | | |
| $1/\sigma_{rev}$ | Time for NRTI and NNRTI resistance to revert back to wild-type (months) | 125 | [16] | | |
| $\alpha_{NRTI \to DTG}$ | Impact of NRTI resistance on DTG resistance mutation acquisition rate | HR=4 | [10] | | |
| *Out of care parameters* | | | | | |
| $1/\gamma_{F_{DTG} \to O}$ | Time from "DTG Failed" to "Out of care" (months) | 110 | [8] | | |
| $1/\gamma_{O \to T_{DTG}}$ | Time from "Out of care" to "DTG Treated" (months) | 22.8 | [9] | | |
| *Druglevel parameter* | | | | | |
| $\rho_{DTG_{detect}}$ | Proportion with detectable drug levels on failing DTG-based ART | 0.626 | [30] | | |
| *Other parameters* | | | | | |
| $\nu_{0,0}$ | probability that a male infects a male (per act) | 0.8% | [31] | | |
| $\nu_{0,1}$ | probability that a male infects a female (per act) | 0.3% | [31] | | |
| $\nu_{1,0}$ | probability that a female infects a male (per act) | 0.3% | [31] | | |
| $\rho_{0,0}$ | percentage of MSM | 5% | [32] | | |
| $\tilde{\mu}^i$ | relative mortality risk | | [33, 34] | | |
| | (Ref: suppressed indiv. with CD4>500) | | | CD4 class | |
| | | 1 | 2 | 3 | 4 |
| | $\tilde{\mu}^i_{I/D}$: not treated ($I$ and $D$) | 1.6 | 2 | 4.6 | 40.9-134.4 |
| | $\tilde{\mu}^i_{T_{NNRTI}/T_{PI}}$: started treatment ($T_{NNRTI}$ and $T_{PI}$) | 2.5 | 2.6 | 3.1 | 10-50.7 |
| | $\tilde{\mu}^i_{S_{NNRTI}/S_{PI}}$: suppressed ($S_{NNRTI}$ and $S_{PI}$) | 1 | 1.3 | 2 | 8.3-41.7 |
| | $\tilde{\mu}^i_{F_{NNRTI}/F_{PI}/O}$: failed ($F_{NNRTI}$ and $F_{PI}$) | 3.9 | 3.9 | 4.3 | 11.8-59.7 |

Note: The rates $\sigma_{res}^{NNRTI}$, $\sigma_{res}^{NRTI}$, and $\sigma_{rev}$ are included in the resistance matrices, see table 4.

### 3 Model simulation

#### 3.1 Default scenario and uncertainty range

Model simulations are performed using the resistance-related parameters outlined in Table 7. Additionally, we conduct simulations using parameter values adjusted to either favor or impede the emergence of resistance to derive an uncertainty range (see Box 1) that complements the main simulation.

Box 1| Uncertainty range

In our simulations, the choice for dolutegravir resistance-relevant parameters is difficult as there is little data available. We therefore model - in addition to our main analysis - a parameter set with more pessimistic, and more optimistic assumptions. The outcomes will then be interpreted as *Uncertainty Range*. This range is **not** to be interpreted as confidence interval, but represents scenarios with plausible parameterizations making conservative or liberal assumptions. (For a full assessment of possible outcomes, see section 8.)

**Table 7:** Main parameters in the prospective scenarios.

| Parameter | Definition | Default | Pessimistic resistance parameters | Optimistic resistance parameters |
|---|---|---|---|---|
| $\rho_{DTG_{detect}}$ | Proportion with detectable drug levels on failing DTG-based ART | 0.626 | 0.814 | 0.482 |
| $R_{DTG}$ | DTG DRM reversion rate, see section 2.3 | 2 Years | 3 Years | 2 Years |
| $\alpha_{NRTI \to DTG}$ | Impact of NRTI resistance on DTG resistance mutation acquisition rates | HR=4 | HR=5 | HR=3 |
| $E_{DTG \to DTG}$ | Impact of DTG resistance on DTG efficacy | 3.24 | 3.9 | 2.7 |
| $T_{DTG}$ | Transmission probability of DTG resistance mutations compared to NNRTI resistance | Same | Same | -20% |

#### 3.2 Prospective scenarios

We simulate the HIV epidemic in South Africa up to 2040 with DTG used as initial first-line regimen (for ART-initiators), and patients on NNRTI-based regimens being switched to a DTG-based regimen starting in 2020. The modelled number of people living with HIV increases at a slower rate than before 2020, and the number of new HIV infections continues to decrease, as well as the number of AIDS-related deaths (figure 4 A-C).
The modelled HIV epidemic in South Africa continues the trends in reaching the UNAIDS goals (figure 4 D-F).

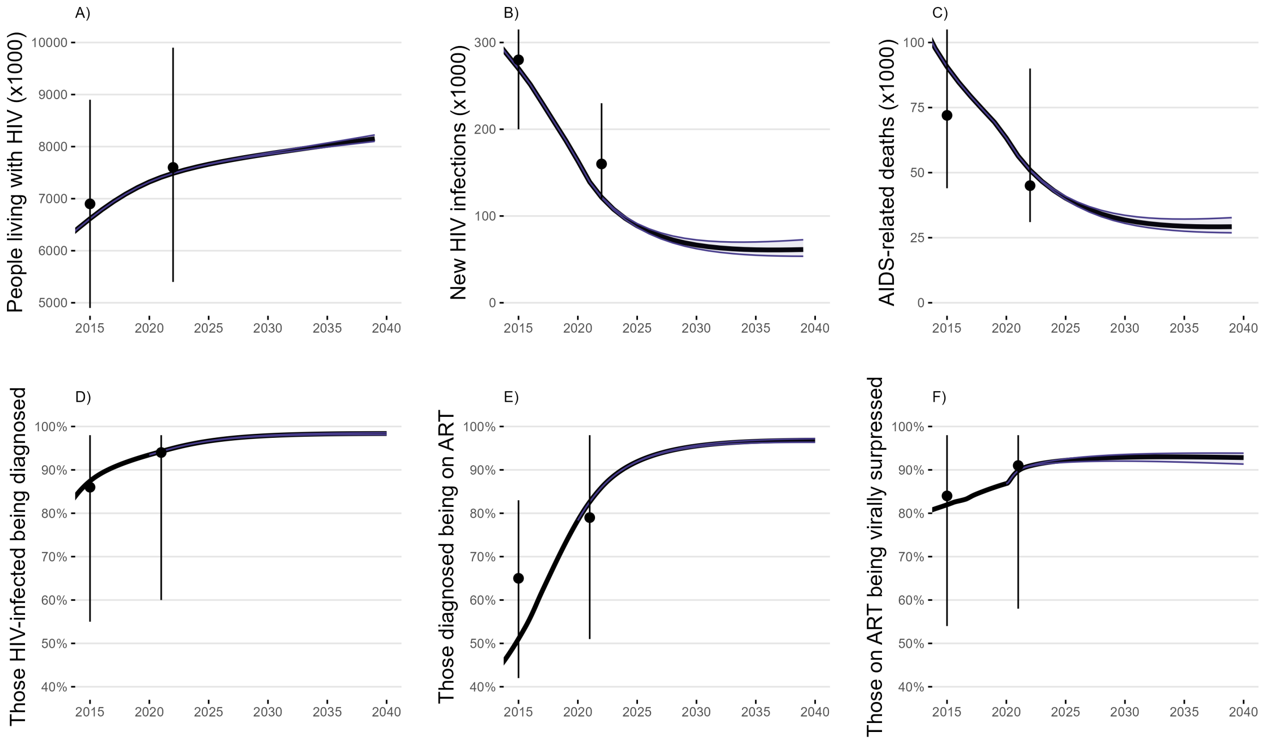

**Figure 4:** Modelled HIV epidemic in South Africa from 2020 to 2040. A) Number of people living with HIV. B) Number of new HIV infections. C) Number of AIDS-related deaths. D) UNAIDS goal 1: Proportion of people living with HIV being diagnosed. E) UNAIDS goal 2: Proportion of people with HIV diagnosis being on ART. F) UNAIDS goal 3: Proportion of people on ART being virally suppressed. Points and errorbars correspond to UNAIDS estimates; shaded area corresponds to the uncertainty range (see box 1 and table 7).

After the switch to DTG-based ART, the number of people on DTG-based ART is rapidly increasing, transitioning to a modest but steady increase in 2023. The number of individuals with viremia on DTG-based ART remains stable in this phase. The population CD4 levels continue to improve in those on DTG-based ART (figure 5).

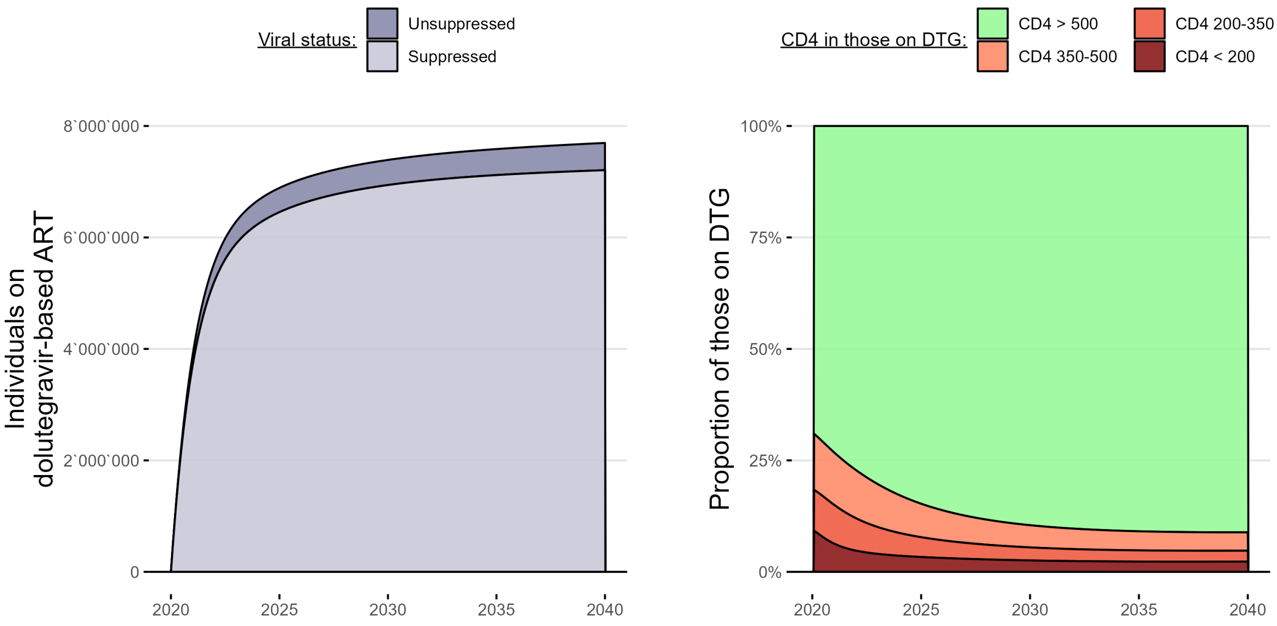

**Figure 5:** Modelled Viral load status and CD4 levels of individuals on DTG-based ART.

#### 3.3 Counterfactual scenario: DTG resistance mitigation strategies

We use counterfactual scenarios to evaluate strategies to mitigate acquired and transmitted DTG resistance such as those proposed in the ongoing RESOLVE study[35]. The following strategies are investigated:

1. **Baseline scenario:**
   Virologic failure on DTG-based ART is managed according to the current treatment guidelines in South Africa, where PI-based ART coverage in South Africa is relatively low at 4%[6], and switching to PI-based ART is only restrictively included in HIV management guidelines. Individuals on failing DTG-based ART remain viremic for extended periods of time.

2. **Switch to PI-based ART:**
   Individuals on failing DTG-based ART are immediately switched to PI-based ART once treatment failure has been detected. Immediate switching can only be done after detecting the virologic failure, i.e., depends on the frequency of viral load monitoring. We assess two options: i) switching to PI-based ART is done after an average time of 6 months after turning viremic on DTG-based ART, and ii) switching to PI-based ART is done after an average time of 12 months after turning viremic on DTG-based ART.

3. **GRT-informed switching to PI-based ART:**
   Upon detection of viremia in individuals on DTG-based ART, they undergo druglevel testing, and in case of detectable concentrations followed by genotypic resistance testing. They are then switched to PI-based ART in case of DTG resistance. The time from turning viremic to switching thus depends on two durations: First, on viral load testing frequency for detecting virologic failure, and second on the time taken from detecting virologic failure to performing druglevel- and genotypic resistance testing, receiving results, and having the individual return to implement treatment switch. We here assess four options: i) time to detecting viremia is 6 months, time to switching in case of DTG resistance is 6 months; ii) time to detecting viremia is 12 months, time to switching in case of DTG resistance is 6 months. This resulted in an average range of time on failing DTG-based ART ranging from 1 to 1.5 years.

All counterfactual scenarios involve increased treatment switching for those on DTG-based ART, but differ in who and when individuals are switched to PI-based ART. The number of people on PI-based ART varies across counterfactual scenarios (figure 6). In scenarios with immediate switch to PI-based ART, the number of individuals on PI-based ART by 2040 is similar to those on DTG-based ART.

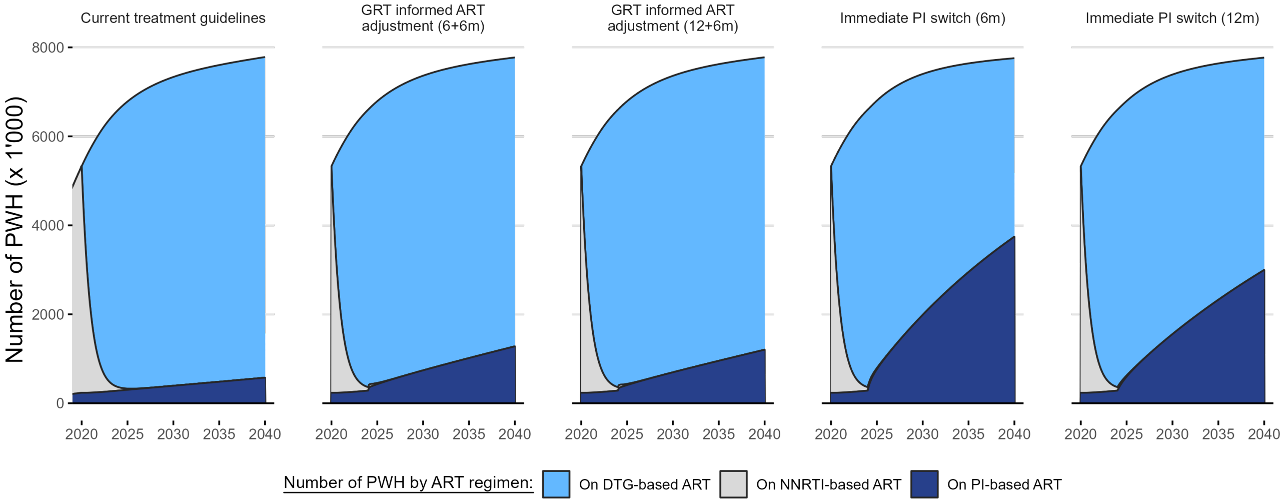

**Figure 6:** Modelled number of individuals by ART regimen for each counterfactual scenario (see section 3.3).

##### 3.4 Sensitivity analyses

In previous works, the sensitivity of parameters related to transmission and NNRTI resistance were thoroughly assessed[1, 2].

We conducted sensitivity analyses by changing one variable at a time while fixing the other parameters for: mutation acquisition rate, reversion rate, NRTI impact on DTG resistance emergence risk, Impact of DTG resistance on DTG efficacy, transmission probability of DTG resistance mutations compared to NNRTI resistance transmission, and the proportion of people with detectable drug levels on failing DTG-based ART, described in table 8 (figure 7).

In addition, we performed a variance-based global sensitivity analysis on these variables. 1'000 bootstrap replicates per included parameter were performed in a Monte Carlo estimation of first oder and total Sobol' indices (see box 2) on population levels of transmitted and acquired DTG resistance (figure 8).

**Box 2| Sobol' Indices**

Sobol' indices are quantitative measures used in sensitivity analyses to assess the importance of input parameters and their interactions on the variability of model outputs. They thereby help identify influential factors in complex models. Sobol' indices are calculated based on decomposition of the total output variance, disregarding interactions (First-Order indices), and including all orders of interactions (Total indices). Sobol' indices are dependent on the ranges of input parameters. [36, 37, 38]

**Table 8:** Parameter ranges used in sensitivity analyses.

| Parameter | Definition | Value | Lower bound | Upper bound |
|---|---|---|---|---|
| $r_{DRM_i}$ | Time to acquire DTG resistance mutation $DRM_i$ based on time on failing ART in the DTG RESIST study (see section 2.3 and equation 1). | 3 Months | 2 Month | 6 Months |
| $R_{DTG}$ | Reversion matrix (for reversion of DTG resistance mutations only), see section 2.3 | 2 Years | 6 Months | 20 Years |
| $\alpha_{NRTI \to DTG}$ | Impact of NRTI resistance on DTG resistance mutation acquisition rates | HR = 4 | HR = 1 | HR = 10 |
| $E_{DTG \to DTG}$ | Impact of DTG resistance on DTG efficacy | 3.24 | 2 | 4 |
| $T_{DTG}$ | Transmission probability of DTG resistance mutations compared to NNRTI resistance | 100% | 5% | 100% |
| $\rho_{DTG_{detect}}$ | Proportion with detectable drug levels on failing DTG-based ART | 0.626 | 0.3 | 1 |

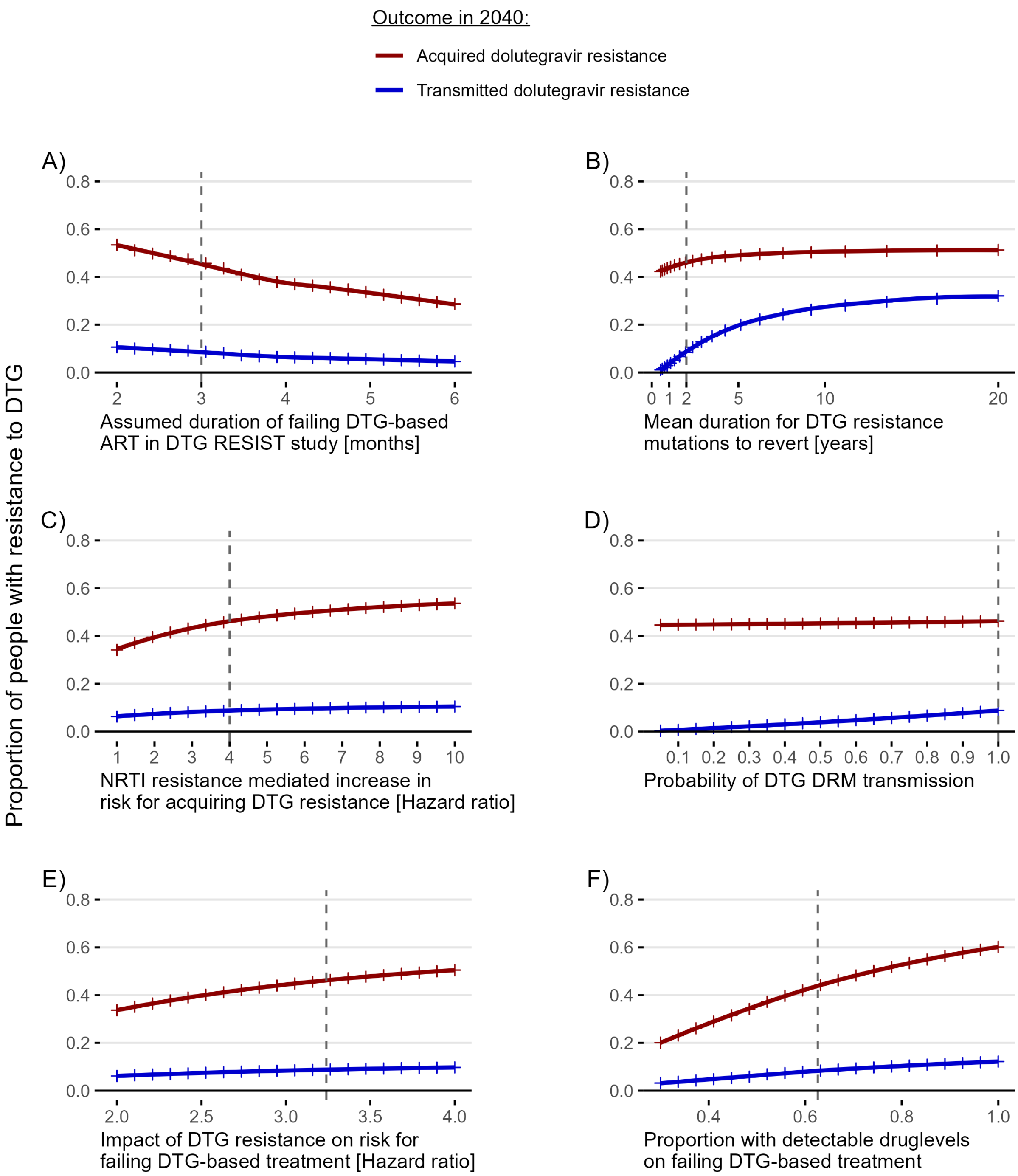

**Figure 7:** Predicted levels of acquired (red) and transmitted (blue) dolutegravir resistance in 2040 were assessed varying only key parameters: A) the acquisition rate of DTG DRMs based on the assumed duration on failing DTG-based ART during which these mutations were acquired, as observed in the DTG RESIST study, B) the time for DTG DRMs to revert given unsuppressed viral replication, C) the impact of NRTI resistance on the risk for acquiring DTG resistance mutations (hazard ratio of NRTI resistance compared to no NRTI resistance), D) the probability of transmitting DTG resistance mutations in a transmission event, E) the impact of DTG resistance on the efficacy of DTG-based ART (hazard ratio of high level DTG resistance compared to no DTG resistance, see section 2.5), F) the proportion of people with detectable drug levels on failing DTG-based ART. Discrete datapoints were modeled (crosses); the line represents a spline over modelled outcomes. Parameter values in the main analysis are represented as gray dotted lines.

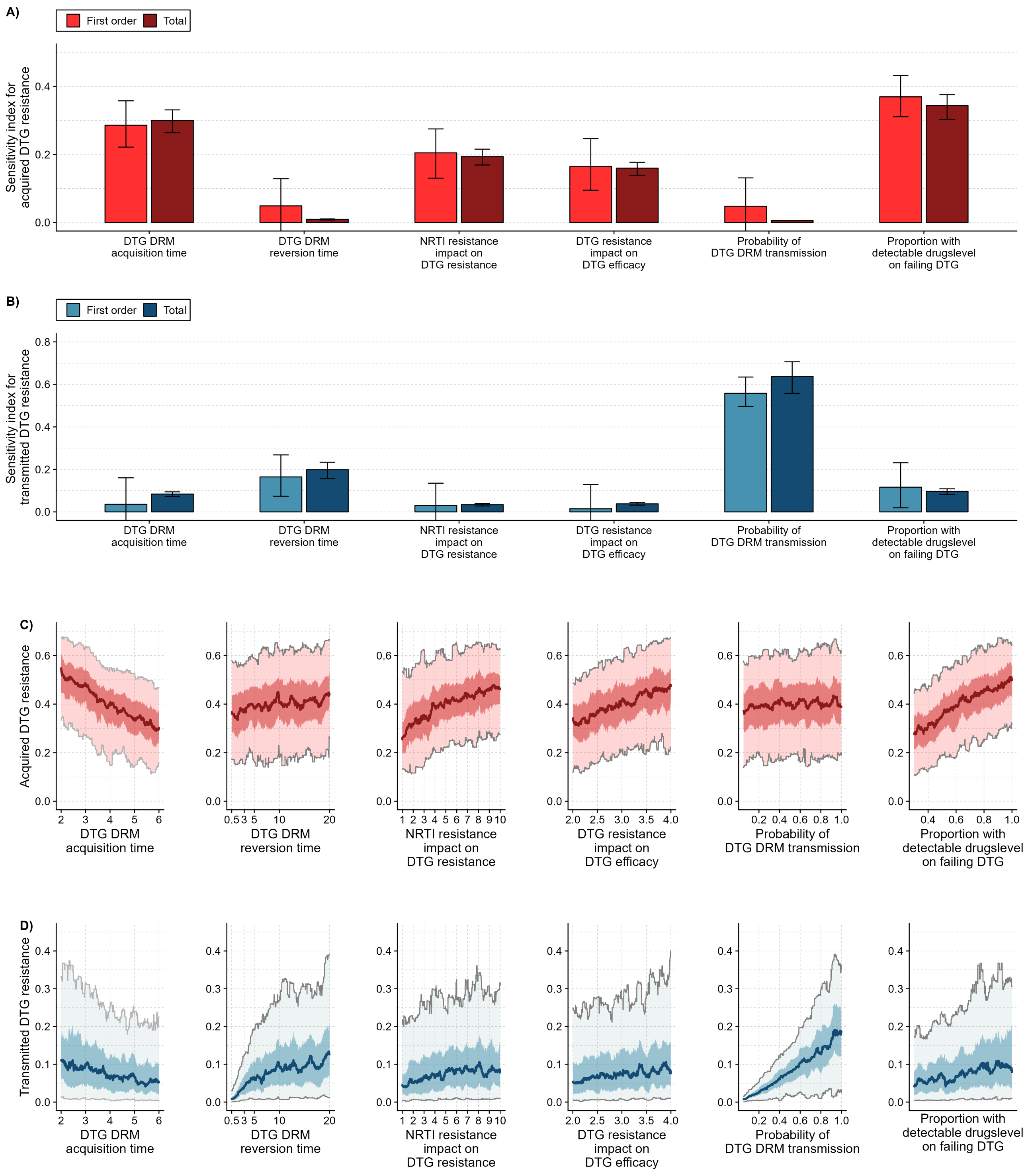

**Figure 8:** Monte Carlo estimation of first- order and total Sobol' indices were implemented using 1,000 samples per included variable. Six key parameters for predicting dolutegravir resistance levels were included: mutation acquisition rate, which are derived from the assumed time people in the DTG RESIST study were on failing dolutegravir-based ART (ranging from 2 to 6 months, see eq. 1); dolutegravir DRM reversion rates (range from 0.5 to 20 years); Impact of NRTI resistance on the risk for acquiring dolutegravir DRMs (hazard ratio for NRTI resistance compared to no resistance ranging from 1, no increased risk, to 10 times higher risk); assumed impact of dolutegravir resistance on dolutegravir efficacy (ranging from a hazard ratio of 2 to 4 for high level DTG resistance compared to no DTG resistance); transmission probability of dolutegravir DRMs compared to NNRTI resistance transmission (ranging from 0.05, dolutegravir DRMs are 20 times less likely to be transmitted compared to NNRTI resistance, to 1, dolutegravir DRMs are transmitted with the same probability as NNRTI resistance); proportion of people with detectable druglevel on failing DTG-based ART (ranging from 30% to 100%). First-order and total Sobol' sensitivity indexes for A) acquired and B) transmitted dolutegravir resistance. Outcomes (proportion with DTG resistance) in 2040 in C) acquired dolutegravir resistance, and in D) transmitted dolutegravir resistance. Color bands represent from light to dark 95%, 50%, and median of realized outcomes of all samples using a rolling window with width corresponding to 5% of the data.

### 4 Model ODEs

#### 4.1 Description of the compartments

Table 9 describes the compartments used in the model, while model ODEs are given in Equations 6.

**Table 9:** Description of the compartments used in the model.

| Notation | Description | Definition |
|---|---|---|
| *Dimensions/Compartments* | | |
| $j$ | index for the 2nd dimension (CD4 counts) | $j = 1, 2, 3, 4$ (4 CD4 strata) |
| $k$ | index for the 3rd dimension (gender) | $k = 0$: men, $k = 1$: women |
| $l$ | index for the 4th dimension (Resistance-Genotype) | $l = 1, ..., g_{tot}$ with $g_{tot} = 2^m$ (m = Number of Mutations) |
| $I^{jkl}(t)$ | number of infected (not diagnosed) indiv. | |
| $D_{jkl(t)},$ | number of diagnosed (not treated) indiv. | |
| $O^{jkl}(t)$ | number of out of care indiv. | |
| *NNRTI-based treatment* | | |
| $T^{jkl}_{NNRTI}(t),$ | number of indiv. that have started NNRTI-based treatment for less than 3 months | |
| $S^{jkl}_{NNRTI}(t),$ | number of suppressed indiv. on NNRTI-based treatment | |
| $F^{jkl}_{NNRTI}(t),$ | number of indiv. failing NNRTI-based treatment | |
| *PI-based treatment* | | |
| $T^{jkl}_{PI}(t)$ | number of indiv. that have started PI-based treatment for less than 3 months | |
| $S^{jkl}_{PI}(t)$ | number of suppressed indiv. on PI-based treatment | |
| $F^{jkl}_{PI}(t)$ | number of indiv. failing PI-based treatment | |
| *DTG-based treatment* | | |
| $T^{jkl}_{DTG}(t)$ | number of indiv. that have started DTG-based treatment for less than 3 months | |
| $S^{jkl}_{DTG}(t)$ | number of suppressed indiv. on DTG-based treatment | |
| $F^{jkl}_{recent}(t)$ | number of indiv. failing DTG-based treatment, on avg. 6 months | |
| $F^{jkl}_{int}(t)$ | number of indiv. failing DTG-based treatment, on avg. between 6 months and 1.5 years | |
| $F^{jkl}_{long}(t)$ | number of indiv. failing DTG-based treatment, on avg. over 1.5 years | |
| *Aggregated compartments* | | |
| $Susc^k$ | number of susceptible indiv. of gender $k$ | |
| $Inf^{kl}_u(t)$ | number of undiagnosed indiv. | $Inf^{kl}_u(t) := I^{kl}(t)$ |
| $Inf^{kl}_d(t)$ | number of infectious diagnosed indiv. | |

#### 4.2 Model ODEs

The rates $\gamma$ represent transition between care stages, $\nu_{CD4}$ the transition between CD4 stages and $\mu_{ij}$ the mortality. The rate $\sigma_{rev}$ represents reversion of NNRTI-resistance when no more drug pressure is exerted, while $\sigma_{res}^{NNRTI}$ and $\sigma_{res}^{NRTI}$ represents the rates of acquiring NNRTI-resistance and NRTI-resistance, respectively, when an individual is failing NNRTI-based treatment. To model new infections, we use $\beta_u$ and $\beta_d$ the respective monthly number of sexual contacts among undiagnosed and diagnosed individuals, $\rho_{k,k}$ the assumed proportion of heterosexual individuals within men and women and $\nu_{k,k'}$ the probability of HIV transmission per sexual act.

$$
\begin{aligned}
\dot{I}^{jkl}(t) =& + \beta_u \left( \sum_{x=1}^{l} \rho_{1-k,k}\nu_{1-k,k} \frac{Susc_k}{N_k} Inf_u^{(1-k)l} \cdot Tr_{l,x} + \sum_{x=1}^{l} \rho_{k,k}\nu_{k,k} \frac{Susc_k}{N_k} Inf_u^{kl} \cdot Tr_{l,x} \right) \mathbb{1}_{j=1} \\
& + \beta_d \left( \sum_{x=1}^{l} \rho_{1-k,k}\nu_{1-k,k} \frac{Susc_k}{N_k} Inf_d^{(1-k)l} \cdot Tr_{l,x} + \sum_{x=1}^{l} \rho_{k,k}\nu_{k,k} \frac{Susc_k}{N_k} Inf_d^{kl} \cdot Tr_{l,x} \right) \mathbb{1}_{j=1} \\
& - \nu_{CD4}^{I,j} \cdot I^{jkl}(t)\mathbb{1}_{j\leq 3} + \nu_{CD4}^{I,j-1} \cdot I^{(j-1)kl}(t)\mathbb{1}_{j\geq 2} \\
& - \sum_{x=1}^{l} R_{x,l} \cdot I^{jkl}(t) + \sum_{x=l}^{g_{tot}} R_{l,x} \cdot I^{jkx}(t) \\
& - \gamma_{I\to D}^{jk}(t) \cdot I^{jkl}(t) \\
& - \mu_I^j \cdot I^{jkl}(t),
\end{aligned}
$$

$$
\begin{aligned}
\dot{D}^{jkl}(t) =& + \gamma_{I\to D}^{jk}(t) \cdot I^{jkl}(t) \\
& - \nu_{CD4}^{D,j} \cdot D^{jkl}(t)\mathbb{1}_{j\leq 3} + \nu_{CD4}^{D,j} \cdot D^{(j-1)kl}(t)\mathbb{1}_{j\geq 2} \\
& - \sum_{x=1}^{l} R_{x,l} \cdot D^{jkl}(t) + \sum_{x=l}^{g_{tot}} R_{l,x} \cdot D^{jkx} \\
& - (\gamma_{D\to T_{NNRTI}}^{j}(t) + \gamma_{D\to T_{DTG}}^{jk}(t)) \cdot D^{jkl}(t) \\
& - \mu_D^j \cdot D^{jkl}(t),
\end{aligned}
$$

$$
\begin{aligned}
\dot{O}^{jkl}(t) =& + \gamma_{F_{DTG}\to O} \cdot (F_{recent}^{jkl}(t) + F_{int}^{jkl}(t) + F_{long}^{jkl}(t)) \\
& + \nu_{CD4}^{F_{NNRTI},j-1} \cdot O^{(j-1)kl}(t)\mathbb{1}_{j\geq 2} - \nu_{CD4}^{F_{NNRTI},j} \cdot O^{jkl}(t)\mathbb{1}_{j\leq 3} \\
& - \sum_{x=1}^{l} R_{x,l} \cdot O^{jkl}(t) + \sum_{x=l}^{g_{tot}} R_{l,x} \cdot O^{jkx}(t) \\
& - \gamma_{O\to T_{DTG}} \cdot O^{jkl}(t) \\
& - \mu_{F_{NNRTI}}^j \cdot O^{jkl}(t)
\end{aligned}
$$

$$
\begin{aligned}
\dot{T}_{NNRTI}^{jkl}(t) =& + \gamma_{D\to T_{NNRTI}}^{jk}(t) \cdot D^{jkl}(t) \\
& + \left( \nu_{CD4}^{T_{NNRTI},j-1} \cdot T_{\substack{(j-1)kl \\ NNRTI}}(t) - \tilde{\nu}_{CD4}^{T_{NNRTI},j-1} \cdot T_{NNRTI}^{jkl}(t) \right) \mathbb{1}_{j\geq 2} \\
& + \left( \tilde{\nu}_{CD4}^{T_{NNRTI},j} \cdot T_{NNRTI}^{(j+1)kl}(t) - \nu_{CD4}^{T_{NNRTI},j} \cdot T_{NNRTI}^{jkl}(t) \right) \mathbb{1}_{j\leq 3} \\
& - (\gamma_{T_{NNRTI}\to S_{NNRTI}}^{jl} + \gamma_{T_{NNRTI}\to F_{NNRTI}}^{jl}) \cdot T_{NNRTI}^{jkl}(t) \\
& - \mu_{T_{NNRTI}}^j \cdot T_{NNRTI}^{jkl}(t)
\end{aligned}
$$

$$
\begin{aligned}
\dot{S}^{jkl}_{NNRTI}(t) = &+ \gamma^{jl}_{T_{NNRTI}\to S_{NNRTI}} \cdot T^{jkl}_{NNRTI}(t) \\
&+ \gamma^{jl}_{F_{NNRTI}\to S_{NNRTI}} \cdot F^{jkl}_{NNRTI}(t) \\
&- \tilde{\nu}^{S_{NNRTI},j-1}_{CD4} \cdot S^{jkl}_{NNRTI}(t)\mathbb{1}_{j\geq 2} + \tilde{\nu}^{S_{NNRTI},j}_{CD4} \cdot S^{(j+1)kl}_{NNRTI}(t)\mathbb{1}_{j\leq 3} \\
&- (\gamma^{jl}_{S_{NNRTI}\to F_{NNRTI}} + \gamma_{S_{NNRTI}\to S_{DTG}}(t)) \cdot S^{jkl}_{NNRTI}(t) \\
&- \mu^{j}_{S_{NNRTI}} \cdot S^{jkl}_{NNRTI}(t)
\end{aligned}
$$

$$
\begin{aligned}
\dot{F}^{jkl}_{NNRTI}(t) = &+ \gamma^{jl}_{S_{NNRTI}\to F_{NNRTI}} \cdot S^{jkl}_{NNRTI}(t) \\
&+ \gamma^{jl}_{T_{NNRTI}\to F_{NNRTI}} \cdot T^{jkl}_{NNRTI}(t) \\
&+ \nu^{F_{NNRTI},j-1}_{CD4} \cdot F^{(j-1)kl}_{NNRTI}(t)\mathbb{1}_{j\geq 2} - \nu^{F_{NNRTI},j}_{CD4} \cdot F^{jkl}_{NNRTI}(t)\mathbb{1}_{j\leq 3} \\
&- \sum_{x\in P_{NNRTI}} A_{l,x} \cdot F^{jkl}_{NNRTI}(t) + \sum_{x\in P_{NNRTI}} A_{x,l} \cdot F^{jkx}_{1,elig}(t) \\
&- \sum_{x\notin P_{NNRTI}} R_{l,x} \cdot F^{jkl)}_{NNRTI}(t) + \sum_{x\notin P_{NNRTI}} R_{x,l} \cdot F^{jkx}_{NNRTI}(t) \\
&- (\gamma^{jl}_{F_{NNRTI}\to S_{NNRTI}} + \gamma^{j}_{F_{NNRTI}\to T_{PI}}(t) + \gamma^{j}_{F_{NNRTI}\to T_{DTG}}(t)) \cdot F^{jkl}_{NNRTI}(t) \\
&- \mu^{j}_{F_{NNRTI}} \cdot F^{jkl}_{NNRTI}(t),
\end{aligned}
$$

$$
\begin{aligned}
\dot{T}^{jkl}_{PI}(t) = &+ \gamma^{j}_{F_{NNRTI}\to T_{PI}}(t) \cdot F^{jkl}_{NNRTI}(t) \\
&+ \gamma^{j}_{F_{DTG}\to T_{PI}}(t) \cdot (F^{jkl}_{recent}(t) + F^{jkl}_{int}(t) + F^{jkl}_{long}(t)) \\
&+ \left(\nu^{T_{PI},j-1}_{CD4} \cdot T^{(j-1)kl}_{PI}(t) - \tilde{\nu}^{T_{PI},j-1}_{CD4} \cdot T^{jkl}_{PI}(t)\right)\mathbb{1}_{j\geq 2} \\
&+ \left(\tilde{\nu}^{T_{PI},j}_{CD4} \cdot T^{(j+1)kl}_{PI}(t) - \nu^{T_{PI},j}_{CD4} \cdot T^{jkl}_{PI}(t)\right)\mathbb{1}_{j\leq 3} \\
&- (\gamma^{j}_{T_{PI}\to S_{PI}} + \gamma^{j}_{T_{PI}\to F_{PI}}) \cdot T^{jkl}_{PI}(t) \\
&- \mu^{j}_{T_{PI}} \cdot T^{jkl}_{PI}(t),
\end{aligned}
$$

$$
\begin{aligned}
\dot{S}^{jkl}_{PI}(t) = &+ \gamma^{j}_{T_{PI}\to S_{PI}} \cdot T^{jkl}_{PI}(t) \\
&+ \gamma^{j}_{F_{PI}\to S_{PI}} \cdot F^{jkl}_{PI}(t) \\
&- \tilde{\nu}^{S_{PI},j-1}_{CD4} \cdot S^{jkl}_{PI}(t)\mathbb{1}_{j\geq 2} + \tilde{\nu}^{S_{PI},j}_{CD4} \cdot S^{(j+1)kl}_{PI}(t)\mathbb{1}_{j\leq 3} \\
&- \gamma^{j}_{S_{PI}\to F_{PI}} \cdot S^{jkl}_{PI}(t) \\
&- \mu^{j}_{S_{PI}} \cdot S^{jkl}_{PI}(t),
\end{aligned}
$$

$$
\begin{aligned}
\dot{F}^{jkl}_{PI}(t) = &+ \gamma^{j}_{S_{PI}\to F_{PI}} \cdot S^{jkl}_{PI}(t) \\
&+ \gamma^{j}_{T_{PI}\to F_{PI}} \cdot T^{jkl}_{PI}(t) \\
&+ \nu^{F_{PI},j-1}_{CD4} \cdot F^{(j-1)kl}_{PI}(t)\mathbb{1}_{j\geq 2} - \nu^{F_{PI},j}_{CD4} \cdot F^{jkl}_{PI}(t)\mathbb{1}_{j\leq 3}
\end{aligned}
$$

$$
\begin{aligned}
&- \sum_{x \in P_{PI}} A_{l,x} \cdot F_{PI}^{jkx}(t) + \sum_{x \in P_{PI}} A_{x,l} \cdot F_{PI}^{jkl}(t) \\
&- \sum_{x \notin P_{PI}} R_{x,l} \cdot F_{PI}^{jkx}(t) + \sum_{x \notin P_{PI}} R_{l,x} \cdot F_{PI}^{jkl}(t) \\
&- \gamma^{j}_{F_{PI} \to S_{PI}} \cdot F_{PI}^{jkl}(t) \\
&- \mu^{j}_{F_{PI}} \cdot F_{PI}^{jkl}(t),
\end{aligned}
$$

$$
\begin{aligned}
\dot{T}^{jkl}_{DTG}(t) = &+ \gamma^{jk}_{D \to T_{DTG}}(t) \cdot D^{jkl}(t) \\
&+ \gamma^{j}_{F_{NNRTI} \to T_{DTG}}(t) \cdot F^{jkl}_{NNRTI}(t) \\
&+ \gamma_{O \to T_{DTG}} \cdot O^{jkl}(t) \\
&+ \left( \nu_{CD4}^{T_{NNRTI},j-1} \cdot T_{DTG}^{(j-1)kl}(t) - \tilde{\nu}_{CD4}^{T_{NNRTI},j-1} \cdot T_{DTG}^{jkl}(t) \right) \mathbb{1}_{j \geq 2} \\
&+ \left( \tilde{\nu}_{CD4}^{T_{NNRTI},j} \cdot T_{DTG}^{(j+1)kl}(t) - \nu_{CD4}^{T_{NNRTI},j} \cdot T_{DTG}^{jkl}(t) \right) \mathbb{1}_{j \leq 3} \\
&- (\gamma^{jl}_{T_{DTG} \to S_{DTG}} + \gamma^{jl}_{T_{DTG} \to F_{DTG}}) \cdot T^{jkl}_{DTG}(t) \\
&- \mu^{j}_{T_{NNRTI}} \cdot T^{jkl}_{DTG}(t),
\end{aligned}
$$

$$
\begin{aligned}
\dot{S}^{jkl}_{DTG}(t) = &+ \gamma^{jk}_{T_{DTG} \to S_{DTG}} \cdot T^{jkl}_{DTG}(t) \\
&+ \gamma_{S_{NNRTI} \to S_{DTG}}(t) \cdot S^{jkl}_{NNRTI}(t) \\
&+ \gamma^{jk}_{F_{DTG} \to S_{DTG}} \cdot (F^{jkl}_{recent}(t) + F^{jkl}_{int}(t) + F^{jkl}_{long}(t)) \\
&- \tilde{\nu}_{CD4}^{S_{NNRTI},j-1} \cdot S^{jkl}_{DTG}(t) \mathbb{1}_{j \geq 2} + \tilde{\nu}_{CD4}^{S_{NNRTI},j} \cdot S^{(j+1)kl}_{DTG}(t) \mathbb{1}_{j \leq 3} \\
&- \gamma^{jk}_{S_{DTG} \to F_{DTG}} \cdot S^{jkl}_{DTG}(t) \\
&- \mu^{j}_{S_{NNRTI}} \cdot S^{jkl}_{DTG}(t),
\end{aligned}
$$

$$
\begin{aligned}
\dot{F}^{jkl}_{recent}(t) = &+ \gamma^{j}_{T_{DTG} \to F_{DTG}} \cdot T^{jkl}_{DTG}(t) \\
&+ \gamma^{j}_{S_{DTG} \to F_{DTG}} \cdot S^{jkl}_{DTG}(t) \\
&+ \nu_{CD4}^{F_{NNRTI},j-1} \cdot F^{(j-1)kl}_{recent}(t) \mathbb{1}_{j \geq 2} - \nu_{CD4}^{F_{NNRTI},j} \cdot F^{jkl}_{recent}(t) \mathbb{1}_{j \leq 3} \\
&- \sum_{x \in P_{DTG}} A_{l,x} \cdot \rho_{DTG_{detect}} \cdot F^{jkx}_{recent}(t) + \sum_{x \in P_{DTG}} A_{x,l} \cdot \rho_{DTG_{detect}} \cdot F^{jkl}_{recent}(t) \\
&- \sum_{x \notin P_{DTG}} R_{x,l} \cdot F^{jkx}_{recent}(t) + \sum_{x \notin P_{DTG}} R_{l,x} \cdot F^{jkl}_{recent}(t) \\
&- (\gamma^{jl}_{F_{DTG} \to S_{DTG}} + \gamma^{j}_{F_{DTG} \to T_{PI}} + \gamma_{F_{DTG} \to O} + \gamma_{F_{DTG_{rec}} \to F_{DTG_{int}}}) \cdot F^{jkl}_{recent}(t) \\
&- \mu^{j}_{F_{NNRTI}} \cdot F^{jkl}_{recent}(t),
\end{aligned}
$$

$$
\begin{aligned}
\dot{F}_{int}^{jkl}(t) = &+ \gamma_{F_{DTG_{rec}} \to F_{DTG_{int}}} \cdot F_{recent}^{jkl}(t) \\
&+ \nu_{CD4}^{F_{NNRTI},j-1} \cdot F_{int}^{(j-1)kl}(t)\mathbb{1}_{j\geq 2} - \nu_{CD4}^{F_{NNRTI},j} \cdot F_{int}^{jkl}(t)\mathbb{1}_{j\leq 3} \\
&- \sum_{x \in P_{DTG}} A_{l,x} \cdot \rho_{DTG_{detect}} \cdot F_{int}^{jkx}(t) + \sum_{x \in P_{DTG}} A_{x,l} \cdot \rho_{DTG_{detect}} \cdot F_{int}^{jkl}(t) \\
&- \sum_{x \notin P_{DTG}} R_{x,l} \cdot F_{int}^{jkx}(t) + \sum_{x \notin P_{DTG}} R_{l,x} \cdot F_{int}^{jkl}(t) \\
&- (\gamma_{F_{DTG} \to S_{DTG}}^{jl} + \gamma_{F_{DTG} \to T_{PI}}^{j} + \gamma_{F_{DTG} \to O} + \gamma_{F_{DTG_{int}} \to F_{DTG_{long}}}) \cdot F_{int}^{jkl}(t) \\
&- \mu_{F_{NNRTI}}^{j} \cdot F_{int}^{jkl}(t),
\end{aligned}
$$

$$
\begin{aligned}
\dot{F}_{long}^{jkl}(t) = &+ \gamma_{F_{DTG_{int}} \to F_{DTG_{long}}} \cdot F_{int}^{jkl}(t) \\
&+ \nu_{CD4}^{F_{NNRTI},j-1} \cdot F_{long}^{(j-1)kl}(t)\mathbb{1}_{j\geq 2} - \nu_{CD4}^{F_{NNRTI},j} \cdot F_{long}^{jkl}(t)\mathbb{1}_{j\leq 3} \\
&- \sum_{x \in P_{DTG}} A_{l,x} \cdot \rho_{DTG_{detect}} \cdot F_{long}^{jkx}(t) + \sum_{x \in P_{DTG}} A_{x,l} \cdot \rho_{DTG_{detect}} \cdot F_{long}^{jkl}(t) \\
&- \sum_{x \notin P_{DTG}} R_{x,l} \cdot F_{long}^{jkx}(t) + \sum_{x \notin P_{DTG}} R_{l,x} \cdot F_{long}^{jkl}(t) \\
&- (\gamma_{F_{DTG} \to S_{DTG}}^{jl} + \gamma_{F_{DTG} \to T_{PI}}^{j} + \gamma_{F_{DTG} \to O}) \cdot F_{long}^{jkl}(t) \\
&- \mu_{F_{NNRTI}}^{j} \cdot F_{long}^{jkl}(t). \qquad (6)
\end{aligned}
$$

[12] Hauser A. Acquired HIV drug resistance mutations on first-line antiretroviral therapy in Southern Africa: Bayesian evidence synthesis;. Available from: https://github.com/anthonyhauser/ADR-meta-analysis.

[32] Anova Health Institute. Rapid Assessment of HIV Prevention, Care and Treatment Programming for MSM in South Africa; 2013.

[33] Maduna PH, Dolan M, Kondlo L, Mabuza H, Dlamini JN, Polis M, et al. Morbidity and Mortality According to Latest CD4+ Cell Count among HIV Positive Individuals in South Africa Who Enrolled in Project Phidisa. PLOS ONE. 2015 apr;10(4):e0121843. Available from: http://www.ncbi.nlm.nih.gov/pubmed/25856495http://www.pubmedcentral.nih.gov/articlerender.fcgi?artid=PMC4391777http://dx.plos.org/10.1371/journal.pone.0121843.

[34] Brennan AT, Maskew M, Sanne I, Fox MP. The interplay between CD4 cell count, viral load suppression and duration of antiretroviral therapy on mortality in a resource-limited setting. Tropical medicine & international health : TM & IH. 2013 may;18(5):619-31. Available from: http://www.ncbi.nlm.nih.gov/pubmed/23419157http://www.pubmedcentral.nih.gov/articlerender.fcgi?artid=PMC3625450.

[35] McCluskey S. A Randomized Clinical Trial to Evaluate Solutions for the Management of Virologic Failure on TLD in Sub-Saharan Africa (RESOLVE). Bethesda (MD): National Library of Medicine (US);. Available from: https://classic.clinicaltrials.gov/ct2/show/NCT05373758.

[36] Sobol IM. Global sensitivity indices for nonlinear mathematical models and their Monte Carlo estimates. Mathematics and Computers in Simulation. 2001;55(1-3):271-80. Available from: https://www.scopus.com/inward/record.uri?eid=2-s2.0-0034926426{&}doi=10.1016{%}2FS0378-4754{%}2800{%}2900270-6{&}partnerID=40{&}md5=18ded60db0f323166ec5945126d26ecb.

[37] Jansen MJW. Analysis of variance designs for model output. Computer Physics Communications. 1999;117(1):35-43. Available from: https://www.scopus.com/inward/record.uri?eid=2-s2.0-0033097907{&}doi=10.1016{%}2FS0010-4655{%}2898{%}2900154-4{&}partnerID=40{&}md5=928585846f0e1bd34d96cfa318d3a6c6.

[38] Saltelli A, Annoni P, Azzini I, Campolongo F, Ratto M, Tarantola S. Variance based sensitivity analysis of model output. Design and estimator for the total sensitivity index. Computer Physics Communications. 2010;181(2):259-70. Available from: `https://www.scopus.com/inward/record.uri?eid=2-s2.0-70450224663{&}doi=10.1016{%}2Fj.cpc.2009.09.018{&}partnerID=40{&}md5=d7fb7ed4df039b7ee2b468e1c9444e65`.
